# Calcium-Sensing Receptor Polymorphisms at rs1801725 are Associated with Increased Risk of Secondary Malignancies

**DOI:** 10.1101/2021.02.24.21252297

**Authors:** Ky’Era V. Actkins, Heather K. Beasley, Annika B. Faucon, Lea K. Davis, Amos M. Sakwe

## Abstract

**Purpose:** Dysregulation of systemic calcium homeostasis during malignancy is common in most patients with high grade tumors. However, it remains unclear whether single nucleotide polymorphisms (SNPs) that alter the sensitivity of the calcium-sensing receptor (CaSR) to circulating calcium are associated with primary and/or secondary neoplasms at specific pathological sites in patients of European and African ancestry.

**Methods:** Multivariable logistic regression models were used to analyze the association of *CASR* SNPs with circulating calcium, parathyroid hormone, vitamin D, and primary and secondary neoplasms.

**Results:** Circulating calcium is associated with an increased risk for breast, prostate, and skin cancers. In patients of European descent, the rs1801725 *CASR* SNP is associated with bone-related cancer phenotypes, deficiency of humoral immunity, and a higher risk of secondary neoplasms in the lungs and bone. Interestingly, circulating calcium levels are higher in homozygous patients for the inactivating *CASR* variant at rs1801725 (TT genotype), and this is associated with a higher risk of secondary malignancies.

**Conclusions:** Breast, prostate, and skin cancer patients with homozygous inactivating variants (TT genotype) at the *CASR* rs1801725 locus have a higher risk of developing secondary neoplastic lesions in the lungs and bone, due in part, to cancer-induced hypercalcemia and/or tumor immune suppression.

## Introduction

The calcium-sensing receptor (CaSR) plays an essential role in systemic calcium homeostasis by sensing slight increases in circulating calcium levels. Activation of the receptor by high calcium modulates intracellular signaling pathways that alter parathyroid hormone (PTH) secretion by parathyroid chief cells and renal calcium handling.^1,2^ Besides its role in calcium homeostasis, the CaSR also plays a significant role in tumor progression and metastasis,^3,4^ chemosensitivity of breast and other neoplasms,^5–7^ and the CaSR is a tumor suppressor in some cancer types, including neuroblastoma and colorectal cancer.^8,9^

During malignancy, calcium homeostasis is progressively disrupted by the secretion of osteolytic factors such as parathyroid hormone-related protein (PTHrP) by tumor cells, which eventually results in cancer-induced hypercalcemia (CIH) or humoral hypercalcemia of malignancy.^10–12^ CIH is common in patients with solid tumors, and this often benign paraneoplastic syndrome is associated with high mortality rates, with a median patient survival of 1-3 months from initial diagnosis.^13,14^ Consistent with these observations, high circulating calcium levels are associated with aggressive breast tumors in premenopausal women and larger breast tumors in postmenopausal women.^15,16^ However, it remains unclear whether the potentially high calcium driven tumor progression is associated with the activity of the CaSR.

Several variants and single nucleotide polymorphisms (SNPs) have been reported in the *CASR* gene.^9,17,18^ Some of these SNPs are inactivating (loss-of-function) and are associated with hypercalcemia syndromes, while others are gain-of-function or activating variants associated with hypocalcemia.^19^ Specifically, missense polymorphisms at rs1801725 (e.g., A986S) and rs1801726 (e.g., Q1011E) in exon 7 of the receptor are differentially associated with calcium in various chronic diseases,^20–22^ some cancers,^23,24^ and various heart diseases.^25^ In several other studies, the A986S CaSR variant at rs1801725 is common among subjects of European ancestry, while the Q1011E variant at rs1801726 is common in individuals of African descent.^20,23,24,26–28^ However, it remains unclear whether the expression of polymorphic *CASR* variants at these loci is associated with circulating calcium, PTH, Vitamin D and whether these factors influence cancer diagnosis at specific pathological sites in patients of European versus African descent.

Our analyses reveal that circulating calcium, PTH, and vitamin D levels are associated with neoplasms at multiple pathological sites, including breast, prostate, and skin cancers. *CASR* polymorphisms at rs1801725, but not at rs1801726, are associated with calcium levels and deficiency of humoral immunity in cancer patients, and an increased risk of secondary neoplastic lesions in the lungs and bone. This study reveals that dysregulation of calcium homeostasis, via expression of CaSR variants at rs1801725, may be an important driver for the progression and metastasis of hypercalcemia prone cancers. Screening of patients for *CASR* variants at this locus may lead to improved management of high calcium driven tumor progression.

## Materials and Methods

### Patient samples

The Vanderbilt University Medical Center (VUMC) maintains a clinical research warehouse, known as the Synthetic Derivative (SD), which includes a de-identified mirror image of the VUMC electronic health records (EHR) available for research purposes.^29^ The SD contains over 3 million records with data on demographics, billing codes according to the International Classification of Diseases (ICD 9th and 10th revisions), procedural codes, clinical notes, medications, and laboratory measurements. In addition, over 250,000 DNA samples are linked to these records in a biorepository (BioVU).

Inclusion criteria for this study were EHRs from individuals with at least 5 ICD codes of any type on separate days over a 3-year period, which defined a “data floor” to select a population of VUMC patients enriched for primary care. ICD9 and ICD10 codes were then mapped to hierarchical phenotype classifications (i.e., “phecodes”) using the phecode map version 1.2 and 1.2b1.^30^ Phecodes were used to identify primary cancer phenotypes at specific pathological sites (e.g., “Cancer of mouth,” “Cancer of the esophagus,” etc.), and secondary malignancies (**Table S1**). Cases were identified by the presence of at least one cancer phecode (which required at least two corresponding ICD codes) and controls were those who did not have any cancer phecodes. Two ancestry-specific datasets were subsequently established (**Table 1**), one composed of individuals of European descent herein designated European dataset (n=53,682), and another containing predominantly individuals of African descent designated African dataset (n=10,777).

**Table 1.** Descriptive statistics for European and African descent datasets.

|  | European Dataset | African Dataset | P |
| --- | --- | --- | --- |
| N | 53680 | 10777 |  |
| Average Age (SD) | 48.74 (22.03) | 37.98 (21.57) | < 0.001 |
| Average BMI (SD) | 27.75 (7.25) | 29.04 (8.65) | < 0.001 |
| **Sex (%)** |  |  | < 0.001 |
| Female | 30801 (57.4) | 6767 (62.8) |  |
| Male | 22879 (42.6) | 4010 (37.2) |  |
| **rs1801725 Alleles (%)** |  |  | < 0.001 |
| 0 | 39939 (74.4) | 9976 (92.6) |  |
| 1 | 12736 (23.7) | 773 (7.2) |  |
| 2 | 1005 (1.9) | 20 (0.2) |  |
| **rs1801726 Alleles (%)** |  |  | < 0.001 |
| 0 | 49733 (92.7) | 7487 (69.5) |  |
| 1 | 3858 (7.2) | 2972 (27.6) |  |
| 2 | 87 (0.2) | 309 (2.9) |  |
| **EHR-Reported Race (%)** |  |  | < 0.001 |
| White | 52461 (97.7) | 285 (2.6) |  |
| Black | 59 (0.1) | 10313 (95.7) |  |
| **Calcium (mg/dL)** |  |  |  |
| N | 51176 | 9878 |  |
| Median [IQR] | 9.30 [9.05, 9.60] | 9.40 [9.10, 9.60] | < 0.001 |
| **Ionized Calcium (mg/dL)** |  |  |  |
| N | 13899 | 2336 |  |
| Median [IQR] | 4.52 [4.32, 4.73] | 4.57 [4.35, 4.80] | < 0.001 |
| **Vitamin D (ng/mL)** |  |  |  |
| N | 18885 | 3341 |  |
| Median [IQR] | 31 [24, 39] | 23 [17, 30.50] | < 0.001 |
| **Parathyroid Hormone (pg/mL)** |  |  |  |
| N | 6937 | 1430 |  |
| Median [IQR] | 56 [36, 91] | 82 [50, 160] | < 0.001 |

Categorical data and allele information were statistically analyzed by chi-square tests, lab data were analyzed by Wilcoxon rank sum tests, and age and BMI were analyzed by Student's t-test. SD = standard deviation, EHR = electronic health record, IQR = Interquartile range.

### Clinical laboratory measurements

All clinical laboratory measurements (labs), including total calcium, ionized calcium, intact parathyrin (parathyroid hormone, PTH), and 1,25 (OH) vitamin D (herein referred to as vitamin D), were extracted from the SD and cleaned using the recently published QualityLab pipeline.^31^ In brief, we were restricted to lab tests for which at least 70% of the observations were measured in the same unit and required that each lab was measured on a minimum of 100 patients resulting in a minimum of 1,000 numeric observations recorded. We then applied lab-specific quality control filters to remove infinite and non-numeric values, as well as observations outside of 4 standard deviations from the overall sample mean, indicative of biologically implausible values due to technical or recording errors, monogenic disorders, or extreme environmental influence. Labs were adjusted for the cubic splines of age (4-knots) and transformed by inverse normalization for analysis by linear and logistic regression models. Median labs in the European and African datasets were then stratified into pre-diagnosis and post-diagnosis values, based on the date of the earliest recorded cancer diagnosis.

### Genotyping and quality control pipeline

Genotyping was performed by using the Illumina MEGA^EX^ array platform.^32^ PLINK v1.9 was used to filter SNPs (< 0.98) and individual subjects (< 0.98) with low call rates, sex discrepancies, and excessive or significantly reduced heterozygosity (|Fhet| > 0.2). Principal component (PC) analysis was performed using flashPCA2 was used to select individuals of European ancestry and, separately, of African ancestry. ^33^ Minor allele frequency was calculated for *CASR* rs1801725 (c.2956G>T, p.Ala986Ser, NM_000388.4) and rs1801726 (c.3031C>G, p.Gln1011Glu, NM_000245.4) using PLINK v1.9.

### Statistical analysis

We first examined the correlation between the clinical labs (circulating calcium, vitamin D, and PTH levels) and cancer status (primary and secondary cancer diagnoses) after accounting for covariates including median age across the medical record, EHR-reported race, and sex. Each lab was then treated as an independent variable in separate multivariable logistic regression models of twenty-two primary and seven secondary cancer diagnoses for a total of 116 models tested.

Next, we tested the relationship between *CASR* SNPs (rs1801725 and rs1801726) and circulating calcium, ionized calcium, vitamin D, and PTH levels after adjusting for sex and the top 10 principal components (PCs) calculated from the genetic data (to adjust for population stratification) in separate multivariable linear regression models. The analysis was performed separately in European and African datasets. A total of 16 regression models were tested. In a subsequent sensitivity analysis, clinical labs were further stratified into pre- and post-cancer diagnoses according to the lab draw date.

Finally, for each SNP, we tested both an additive (rs1801725: GG vs GT vs TT, rs1801726: CC vs CG vs GG) and a recessive (rs1801725: TT vs (GG + GT), rs1801726: GG vs (CC + CG)) genetic model in a logistic regression framework to evaluate the relationship between the *CASR* variants and seven secondary malignant cancers in the European dataset resulting in a total of 14 models tested. Median age across the medical record, sex, and the top 10 PCs were included as covariates in genetic analyses of lab traits and cancer diagnoses.

### Statistical power and correction for multiple comparisons

The primary analyses in this report included a total of 148 independent statistical tests, reflecting *a priori* derived hypotheses. A strict Bonferroni correction of p < 3.38e-04 (0.05/148) was used to determine statistical significance of primary results from four sets of regression analyses (i.e., testing associations between primary cancer and labs, *CASR* SNPs and labs, *CASR* SNPs and primary cancer, and *CASR* SNPs and secondary malignancies). The exploratory phenome-wide association study (PheWAS), a hypothesis *generating* analysis, was not included in the multiple testing correction.

## Results

### Characteristics of European and African datasets

The sample demographics of each of our datasets are shown in **Table 1**. Although the datasets for individuals of European and African descent differed in sample size and average age, both sets had a similar proportion of males and females. The datasets were defined by genetic ancestry clusters mapped onto principal components derived from genetic data. We analyzed the European descent and African descent populations separately in all genetic analyses to avoid population stratification confounders. In the EHR-based data without accompanying genetic data, we did not have access to ancestry information. Previous studies have reported differences in calcium and vitamin D levels by race.^32,33^ Although race is not an appropriate proxy for genetic ancestry, we stratified our EHR-based phenotypic analyses by race to avoid confounding by race.

### Allele frequencies for rs1801725 and rs1801726

Consistent with previous reports,^23^ the A986S variant of the CaSR was more frequent in the European dataset compared to the African dataset. In the European dataset, the minor allele frequency (MAF) for rs1801725 was 0.14 and for rs1801726 was 0.08. In the African dataset, the MAF for rs1801725 was 0.04, while that for rs1801726 was 0.16. The European dataset had more recessive homozygotes for rs1801725 (N_EUR_= 1,005; N_AFR_ = 20) while the African dataset had more recessive homozygotes for rs1801726 (N_AFR_= 309; N_EUR_= 87).^26^

We then carried out a preliminary analysis of the association between rs1801725 and the rs1801726 SNPs and disease phenotypes in these datasets by using additive and recessive PheWAS models. Based on a strict Bonferroni correction (p < 3.65 × 10^−5^), these exploratory analyses revealed that polymorphisms at rs1801725, but not rs1801726, were strongly associated with deficiency of humoral immunity (**Figure S1**). Although we also observed that these *CASR* SNPs were associated with several other disease phenotypes including hypercalcemia and secondary malignancy of respiratory organs, these phenotypes did not pass the strict correction threshold (**Figure S2-4**).

### Circulating calcium, vitamin D, and PTH levels in individuals of European and African ancestries

In a previous study, total serum calcium was found to be higher in African American compared to those in European American subjects,^23^ but the underlying cause for this difference was not further studied. Here, we determined that both circulating total (p = <0.001) and ionized calcium (p = <0.001) levels are higher in individuals of African descent compared to individuals of European descent (**Table 1**). Interestingly, in individuals of African descent, serum vitamin D levels are lower (p = <0.001) while intact PTH levels are higher (p = <0.001) than those in individuals of European descent (**Table 1**). These differences in calciotropic hormone levels suggest that the relatively higher circulating calcium in individuals of African descent may be attributed to reduced vitamin D facilitated reabsorption of calcium in the kidneys and increased PTH mediated bone resorption.

### Association of circulating calcium, vitamin D, and PTH with primary cancer phenotypes

Next, we assessed whether total serum calcium, ionized calcium, vitamin D, and PTH levels are associated with specific primary cancer types (**Figure 1; Tables S2-5)**. This analysis revealed that circulating calcium, vitamin D and PTH levels are independently, but mostly inversely, associated with neoplasms at several pathological sites. However, total serum calcium was positively associated with breast (OR = 1.26, 95% CI = 1.23-1.29, p = 3.83e-91), prostate (OR = 1.18, 95% CI = 1.15-1.21, p = 1.58e-40), and skin (OR = 1.23, 95% CI = 1.21-1.25, p = 4.79e-136) cancers (**Table S2**). Similarly, ionized calcium was positively correlated with skin cancer (OR = 1.07, 95% = 1.04-1.11, p=1.47e-05) and liver cancer (OR = 1.19, 95% = 1.11-1.28, p = 1.55e-06) (**Table S3**), while circulating vitamin D was not only associated with breast (OR = 1.13, 95% = 1.10-1.17, p = 9.89e-14) and skin cancers (OR = 1.06, 95% = 1.03-1.09, p = 2.17e-05), but also thyroid cancer (OR = 1.17, 95% = 1.10-1.25, p = 1.92e-07) (**Table S4**). Meanwhile, PTH was positively associated with cancer of urinary organs (OR = 1.25, 95% = 1.17-1.33, p = 1.16e-11) (**Table S5**).

**Figure 1.**
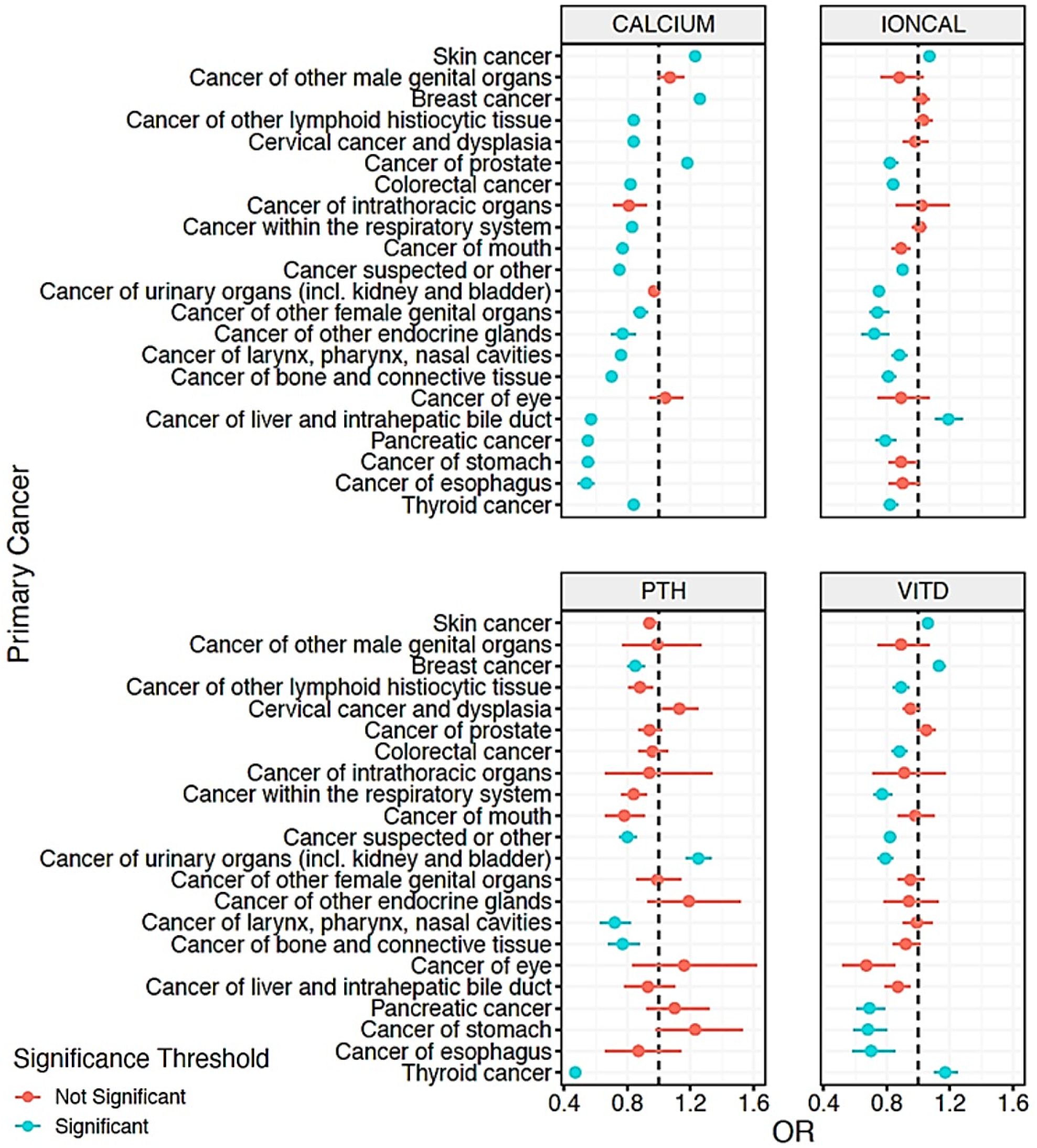
Association of circulating calcium, parathyroid hormone, and vitamin D levels with cancer risk. Forest plots showing the odds associated with circulating levels of the indicated laboratory values and diagnosis of cancer at the indicated pathologic sites. The dashed line in each plot represents an odds ratio (OR) of 1. PTH = intact parathyroid hormone, VITD = vitamin D, IONCAL = ionized calcium.

We further demonstrate that circulating total and ionized calcium are also significantly, but negatively associated with secondary malignancies of lymph nodes, respiratory organs, digestive system, liver, brain/spine, and bone (**Table S6**). Serum vitamin D levels are also significantly negatively associated with secondary malignancies of respiratory organs, digestive system, and liver, while circulating intact PTH levels are significantly associated with secondary malignancy of lymph nodes. Together, this suggests that low circulating calcium and calciotropic hormones are associated with secondary neoplasms at diverse pathological sites.

### Association of CASR SNPS with circulating calcium, vitamin D, and intact PTH

Several studies have reported the association of rs1801725 and rs1801726 *CASR* SNPs with calcium levels.^34^ In our European dataset, we confirmed that polymorphisms at rs1801725, but not rs1801726, were strongly associated with total and ionized calcium and modestly associated with secondary malignancy (p = 0.03). However, the association of these SNPs with Vitamin D, PTH, and primary and secondary malignancies was not statistically significant (**Table 2**). We also found that median calcium values were significantly (p < 0.05) different between all rs1801725 allele carriers (GG vs GT vs TT) and that individuals expressing the TT genotype had the highest median calcium levels compared to those expressing the GG genotype (**Figure 2a, Table S7**). Although this trend was also observed for ionized calcium, only homozygous dominant and heterozygous carriers (GG vs. GT), and homozygous dominant and homozygous recessive carriers (GG vs. TT) had significantly (p < 0.05) different ionized calcium levels (**Figure 2b**). The same analytic approach also revealed a similar trend for individuals of African descent regarding total and ionized serum calcium for carriers of polymorphisms at rs1801725 (GG vs GT) (**Figure 2, Table S8**).

**Table 2.** Relationship between *CASR* exon 7 variants and calcium, PTH, vitamin D and cancer in European descent individuals.

|  | **rs1801725** |  | **rs1801726** |  |
| --- | --- | --- | --- | --- |
|  | **OR (95% CI)** | **p** | **OR (95% CI)** | **p** |
| **Labs** |  |  |  |  |
| Calcium | 1.15 (1.13-1.16) | 8.40E-73* | 0.98 (0.96-1.01) | 0.16 |
| Ionized Calcium | 1.10 (1.07-1.14) | 1.43E-09* | 1.03 (0.97-1.09) | 0.28 |
| Vitamin D | 0.99 (0.96-1.02) | 0.50 | 0.98 (0.94-1.03) | 0.51 |
| Parathyroid Hormone | 1.00 (0.95-1.05) | 0.98 | 1.03 (0.94-1.12) | 0.57 |
| **Primary Cancer** | 1.02 (0.97-1.08) | 0.40 | 1.00 (0.91-1.09) | 0.95 |
| **Secondary Malignancy** | 1.08 (1.01-1.15) | 0.03 | 1.00 (0.88-1.14) | 0.97 |

The additive regression model adjusted for median age, sex, and the first ten principal components was performed between CASR SNPs (rs180125 (G vs T) and rs1801726 (C vs G)) and the indicated laboratory values as well as cancer diagnosis. In this analysis the phecodes used for the primary cancers and secondary malignancies were each collapsed into a single diagnostic code. \* P < 0.001.

**Figure 2.**
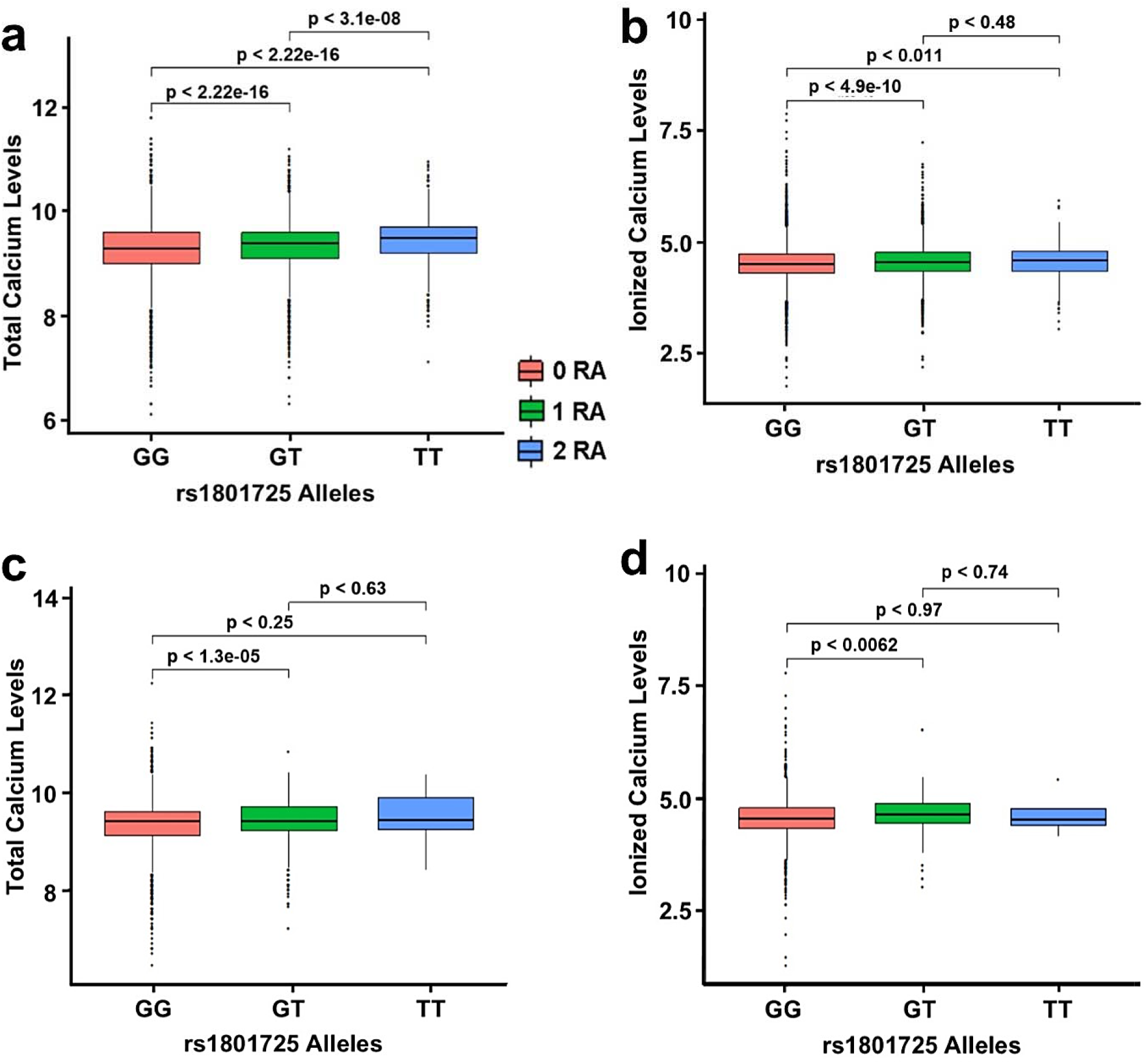
Circulating total calcium and ionized calcium in CaSR rs1801725 allele carriers. The Wilcoxon rank sum test was used to compare circulating total (**a**) and ionized (**b**) calcium levels in individuals of European descent, and total (**c**) and ionized (**d**) calcium levels in individuals of African descent expressing the indicated (color coded) polymorphisms at rs1801725 depicted by the number of T or risk alleles. p < 0.05 was considered statistically significant. RA = risk alleles.

We also confirmed previous reports that the T-allele at rs1801725 is associated with increased total serum calcium and, interestingly, increased ionized calcium levels in the European population (**Table S9**).^23^ The biggest effect was observed with the assessment of total serum calcium levels with an OR of 1.15 (95% CI = 1.13-1.16, p = 8.40e-73), followed by ionized calcium with an OR of 1.10 (95% CI = 1.07-1.14, p = 1.43e-09). A similar linear regression analysis revealed that polymorphisms at rs1801726 in individuals of European descent are not associated with circulating calcium, vitamin D, or PTH levels. We next showed that *CASR* polymorphisms at the rs1801725 locus are associated with circulating total and ionized calcium levels prior to and after cancer diagnosis in individuals of European descent (**Table S10**).

For individuals of African descent, total serum calcium was modestly (p < 0.05) associated with rs1801725 (OR = 1.12, 95% CI = 1.06-1.20, p = 2.34e-04). As in the European population, the rs1801726 locus was not significantly associated with calcium, vitamin D, or PTH (**Table S9**). Together, this suggests that polymorphisms at rs1801725, but not at rs1801726, are associated with circulating serum calcium in individuals of European or African descent.

### Association of CASR SNPs with secondary cancer types

Although assessment of tumor characteristics and/or progression patterns are not available in the EHRs, we hypothesized that the association of CaSR SNPs with tumors at secondary sites (secondary malignancies) is indicative of the progression of primary tumors to these sites. To test this, we first assessed whether the CaSR SNPs were associated with any secondary malignancy (i.e., cases with any secondary malignancy code). This analysis revealed that the polymorphism at rs1801725 (TT genotype), but not rs1801726, was significantly associated (p < 0.05) with secondary malignancies in the European dataset (**Tables S11-12**).

We next determined which secondary cancers were associated with the rs1801725 *CASR* SNP. Using an additive logistic regression model, we observed that the rs1801725-T allele was modestly associated with the presence of secondary malignancy of respiratory organs (OR = 1.21, 95% CI = 1.08-1.35, p = 9.61E-04) and with secondary malignancy of bone (OR of 1.20 (95% CI = 1.06-1.36, p = 4.28e-03) in the European dataset (**Figure 3, Table S11**). Using a recessive model, we confirmed that the TT genotype at rs1801725 was more common among patients with secondary malignancy of bone (OR = 1.82, 95% CI = 1.25-2.65, p = 1.80E-03) (**Table S12**). These results, while supportive of the hypothesis, did not exceed our strict multiple correction threshold. The small sample size of TT allele carriers in the population of African ancestry for rs1801725 precluded further genotype analysis in this population (**Table 1**).

**Figure 3.**
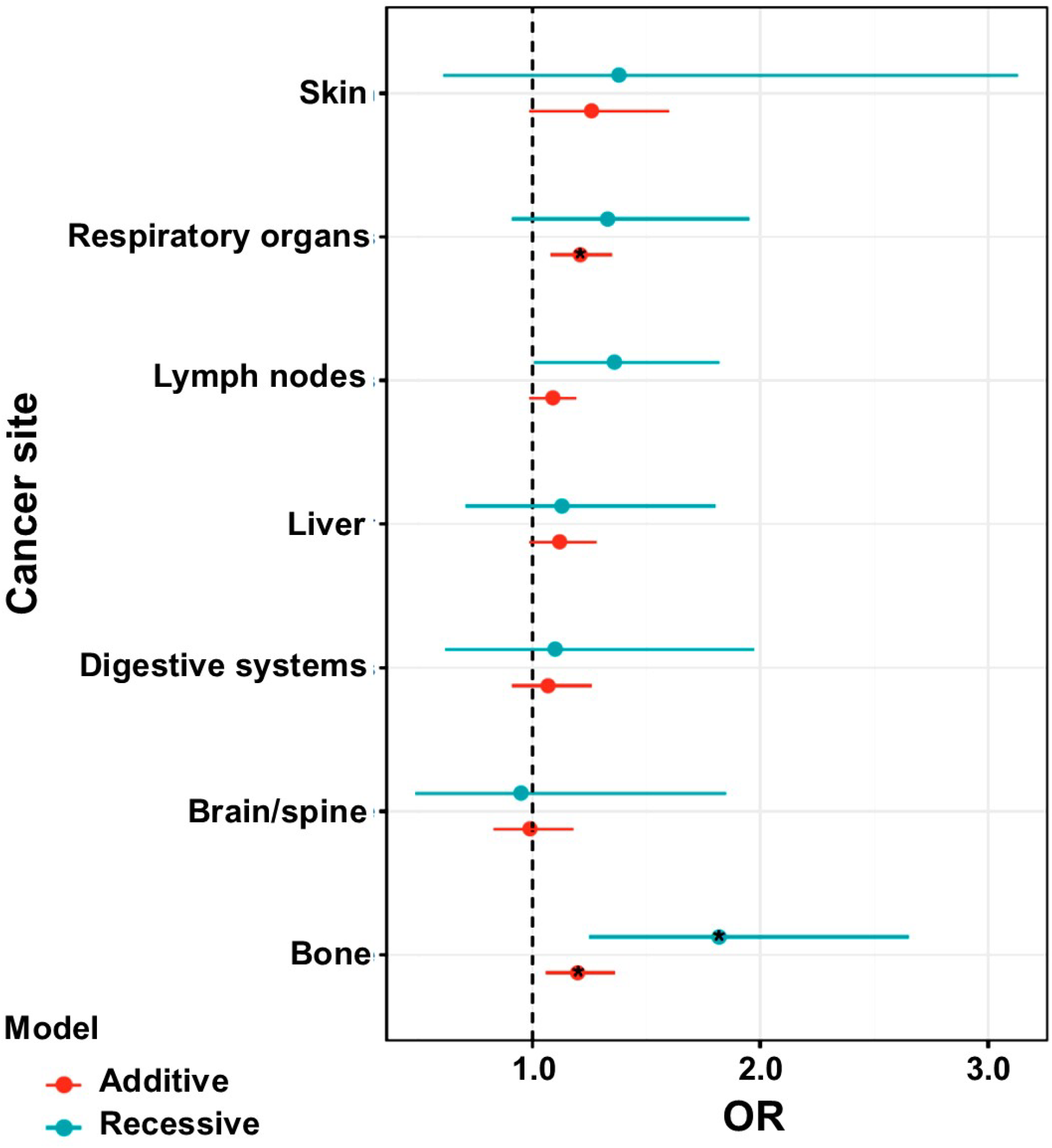
Association of circulating calcium levels with secondary cancer phenotypes. Logistic regression models were performed on secondary cancer and *CASR* variants at rs1801725. The additive (red) and recessive (green) models were adjusted for median age, sex, and the first ten principal components. * indicates P < 0.01 in the logistic regression model. OR = odds ratio.

## Discussion

In this study, we assembled larger, genotyped datasets of individuals of European and African descent. We interrogated whether *CASR* polymorphisms at rs1801725 and rs1801726 are associated with circulating total and ionized calcium, vitamin D, and intact PTH levels, as well as primary cancers and secondary malignancies for individuals with a primary cancer diagnosis. We replicated previous studies showing that circulating calcium levels in individuals of African descent are higher than those of European descent.^23^ Furthermore, we demonstrate that *CASR* variants at rs1801725, but not rs1801726, are associated with calcium and bone-related secondary cancer phenotypes, a higher risk of developing secondary neoplastic lesions in the lungs and bone, and interestingly, deficiency of humoral immunity. Overall, our data suggest that dysregulation of calcium homeostasis via expression of potentially inactivating *CASR* variants at rs1801725 (e.g., A986S) may be an important driver for the progression of primary neoplasms at several pathological sites, including breast, prostate, and skin, as well as metastasis to lung and bone tissues.

This study provides further evidence suggesting that dysregulation of calcium homeostasis is distinct in cancer patients of African and European descent. The distinction is more pronounced in patients expressing polymorphic CaSR variants at rs1801725 rather than at rs1801726. The higher circulating calcium in individuals of African descent is presumably due to lower circulating 1,25-OH vitamin D and higher PTH levels. This is especially interesting as reduced vitamin D levels are associated with reduced reabsorption of calcium in the kidneys,^35^ while higher circulating PTH will lead to increased bone resorption.^36^

This study assembled a relatively large dataset of African American patients comprising 10,777 records from this cohort. We were not able to detect any significant associations between polymorphisms at rs1801725 and circulating calcium or cancer phenotypes in the African descent cohort due to the MAF at this locus. Similar to other studies, rs1801726 was not associated with any of the cancer phenotypes in either the European or the African ancestry datasets.^37^ However, we found modest associations between these *CASR* SNPs in individuals of African descent with several disease phenotypes in our exploratory PheWAS. This includes a positive association between rs1801725 and osteoporosis, whereas previous reports found no associations between osteoporosis and rs1801725 in individuals of European ancestry.^38^ Finally, it is possible that the lack of association between rs1801726 and circulating calcium in the African ancestry sample may suggest a lack of effect of *CASR* variants at this locus on the activity of the receptor. Although some studies have linked the development of hypercalcemia to *CASR* variants at these loci,^22^ direct evidence on the activity, especially with the A986S and the Q1011E receptor variants, remains to be clearly demonstrated. Dysregulation of calcium homeostasis in cancer patients expressing *CASR* variants at rs1801725 may be linked to the reduced activity of the receptor to calcium and therefore, the well documented association between polymorphic variants at this locus and higher circulating calcium.^23,26,28,39^ However, whether the difference in circulating calcium levels and/or the expression of *CASR* variants at rs1801725 underlies the disparate aggressiveness of especially prostate and breast cancers between patients of African and European ancestries remains to be clearly delineated.

Besides hypercalcemia, we also found a significant association between *CASR* rs1801725 polymorphisms and deficiency in humoral immunity in our exploratory analysis. This finding raises the hypothesis that tumor progression and metastasis to secondary sites such as bone may also lead to tumor induced immune suppression. In addition to its role in calcium homeostasis, vitamin D also plays a central role in innate and adaptive immune function through the vitamin D receptor. Individuals predisposed to higher circulating calcium due to vitamin D deficiency may have a weakened immune system that may further increase susceptibility to secondary malignancies. This, coupled with increased PTH and hypercalcemia, may especially compromise high risk populations such as those of African ancestry who disproportionately have higher cancer mortality rates and aggressive tumors.^40^

### Study Limitations

In spite of the robustness of our findings and opportunities for future studies, some limitations of this study are worth noting. First, we were unable to analyze PTHrP, the major osteolytic factor during malignancy. This is because PTHrP levels are not routinely measured in a clinical setting, and the small sample size in our EHR datasets warranted their exclusion during implementation of the QualityLab pipeline. Second, although our dataset containing individuals of African descent was well powered for rs1801725 in the additive models, it was not possible for us to examine the effects of the T-allele in the recessive model because of the MAF within the African descent population. Third, the skewed nature of the frequency of SNPs at rs1801725 or rs1801726 in individuals of African and European ancestry made it difficult to generalize the effects of rs1801725 *CASR* variants in the African population. Fourth, due to differing linkage disequilibrium structures in European and African populations, it is possible that the associations we observed with the European descent do not tag the same causal allele associations in the African descent dataset. Finally, the EHR-based data from which we drew our study population is a clinically ascertained sample, thus, ascertainment biases could impact this study. More functional *in vitro* and *in vivo* studies are needed to understand causal links between CaSR genetic variants and cancer progression and/or metastasis.

In summary, this study provides robust population-level genetic evidence supporting the notion that expression of the inactivating CaSR rs1801725 SNP predisposes breast, prostate, and skin cancer patients to secondary neoplastic lesions in the lung and bone tissues. Whether the development of secondary malignancies at these sites is exclusively mediated by cancer-induced hypercalcemia or includes other complications such as tumor immune suppression remains to be fully elucidated.

## Supporting information

Supplementary PheWAS Data

## Data Availability

The data that support the findings of this study are available from Vanderbilt University Medical Center, but restrictions apply to the availability of these data, which were used under license for the current study, and so are not publicly available. Data are, however, available from the authors upon reasonable request and with permission of Vanderbilt University Medical Center.

## Author Information

Conceptualization: K.V.A., H.K.B., A.B.F., L.K.D., A.M.S.; Data curation: K.V.A., A.B.F. Formal Analysis: K.V.A., A.B.F., L.K.D.; Funding acquisition: L.K.D., A.M.S.; Investigation: K.V.A., A.B.F.; Methodology: K.V.A., A.B.F., L.K.D. Project administration: K.V.A., H.K.B.; Resources: L.K.D.; Supervision: L.K.D., A.M.S.; Visualization: K.V.A.; Writing - original draft: K.V.A., H.K.B., A.B.F.; Writing - reviewing & editing: K.V.A., H.K.B, A.B.F., L.K.D., A.M.S.

## Ethics Declaration

The use of de-identified EHR data for this project was approved by the VUMC Institutional Review Board under IRB #190418. The research was conducted in accordance with the principles of the Helsinki Declaration.

## Notes

### Competing Interest Statement

The authors have declared no competing interest.

### Funding Statement

KVA is funded by the NIH 5T32GM007628 training grant. HKB is funded by the NIH R25 GM059994 and U54 CA163069 grants. AS is supported by SC1 CA211030, P50 CA095103, U54 MD007593. This study used resources from the Advanced Computing Center for Research and Education and the Synthetic Derivative at Vanderbilt University Medical Center which are supported by Shared Instrumentation Grant S10RR025141 and CTSA grants UL1TR002243, UL1TR000445, and UL1RR024975. Genomic data repositories are supported by investigator-led projects that include U01HG004798, R01NS032830, RC2GM092618, P50GM115305, U01HG006378, U19HL065962, R01HD074711; and additional funding sources listed at https://victr.vumc.org/biovu-funding.

## References

1. Tfelt-Hansen J, Brown EM. The calcium-sensing receptor in normal physiology and pathophysiology: a review. Crit Rev Clin Lab Sci. 2005;42(1):35–70. doi:10.1080/10408360590886606

2. Brown EM, Gamba G, Riccardi D, et al. Cloning and characterization of an extracellular Ca(2+)-sensing receptor from bovine parathyroid. Nature. 1993;366(6455):575–580. doi:10.1038/366575a0

3. Kim W, Wysolmerski JJ. Calcium-Sensing Receptor in Breast Physiology and Cancer. Front Physiol. 2016;7:440. doi:10.3389/fphys.2016.00440

4. Masvidal L, Iniesta R, García M, et al. Genetic variants in the promoter region of the calcium-sensing receptor gene are associated with its down-regulation in neuroblastic tumors. Mol Carcinog. 2017;56(4):1281–1289. doi:10.1002/mc.22589

5. Hernández-Bedolla MA, Carretero-Ortega J, Valadez-Sánchez M, Vázquez-Prado J, Reyes-Cruz G. Chemotactic and proangiogenic role of calcium sensing receptor is linked to secretion of multiple cytokines and growth factors in breast cancer MDA-MB-231 cells. Biochimica et Biophysica Acta (BBA) - Molecular Cell Research. 2015;1853(1):166–182. doi:10.1016/j.bbamcr.2014.10.011

6. Liu G, Hu X, Chakrabarty S. Calcium sensing receptor down-regulates malignant cell behavior and promotes chemosensitivity in human breast cancer cells. Cell Calcium. 2009;45(3):216–225. doi:10.1016/j.ceca.2008.10.004

7. Singh N, Aslam MN, Varani J, Chakrabarty S. Induction of calcium sensing receptor in human colon cancer cells by calcium, vitamin D and aquamin: Promotion of a more differentiated, less malignant and indolent phenotype. Mol Carcinog. 2015;54(7):543–553. doi:10.1002/mc.22123

8. Aggarwal A, Prinz-Wohlgenannt M, Tennakoon S, et al. The calcium-sensing receptor: A promising target for prevention of colorectal cancer. Biochim Biophys Acta. 2015;1853(9):2158–2167. doi:10.1016/j.bbamcr.2015.02.011

9. Tennakoon S, Aggarwal A, Kállay E. The calcium-sensing receptor and the hallmarks of cancer. Biochim Biophys Acta. 2016;1863(6 Pt B):1398–1407. doi:10.1016/j.bbamcr.2015.11.017

10. Guise TA, Kozlow WM, Heras-Herzig A, Padalecki SS, Yin JJ, Chirgwin JM. Molecular Mechanisms of Breast Cancer Metastases to Bone. Clinical Breast Cancer. 2005;5:S46–S53. doi:10.3816/cbc.2005.s.004

11. Mundy GR. Metastasis to bone: causes, consequences and therapeutic opportunities. Nat Rev Cancer. 2002;2(8):584–593. doi:10.1038/nrc867

12. Sanders JL, Chattopadhyay N, Kifor O, Yamaguchi T, Butters RR, Brown EM. Extracellular calcium-sensing receptor expression and its potential role in regulating parathyroid hormone-related peptide secretion in human breast cancer cell lines. Endocrinology. 2000;141(12):4357–4364. doi:10.1210/endo.141.12.7849

13. Ralston SH, Gallacher SJ, Patel U, Campbell J, Boyle IT. Cancer-associated hypercalcemia: morbidity and mortality. Clinical experience in 126 treated patients. Ann Intern Med. 1990;112(7):499–504. doi:10.7326/0003-4819-112-7-499

14. Stewart AF. Hypercalcemia Associated with Cancer. N Engl J Med. 2005;352(4):373–379. doi:10.1056/NEJMcp042806

15. Almquist M, Anagnostaki L, Bondeson L, et al. Serum calcium and tumour aggressiveness in breast cancer: a prospective study of 7847 women. Eur J Cancer Prev. 2009;18(5):354–360. doi:10.1097/CEJ.0b013e32832c386f

16. Thaw SSH, Sahmoun A, Schwartz GG. Serum calcium, tumor size, and hormone receptor status in women with untreated breast cancer. Cancer Biol Ther. 2012;13(7):467–471. doi:10.4161/cbt.19606

17. Hendy GN, D’Souza-Li L, Yang B, Canaff L, Cole DE. Mutations of the calcium-sensing receptor (CASR) in familial hypocalciuric hypercalcemia, neonatal severe hyperparathyroidism, and autosomal dominant hypocalcemia. Hum Mutat. 2000;16(4):281–296. doi:10.1002/1098-1004(200010)16:4<281::AID-HUMU1>3.0.CO;2-A

18. Pidasheva S, D’Souza-Li L, Canaff L, Cole DEC, Hendy GN. CASRdb: calcium-sensing receptor locus-specific database for mutations causing familial (benign) hypocalciuric hypercalcemia, neonatal severe hyperparathyroidism, and autosomal dominant hypocalcemia. Hum Mutat. 2004;24(2):107–111. doi:10.1002/humu.20067

19. Thakker RV. Diseases associated with the extracellular calcium-sensing receptor. Cell Calcium. 2004;35(3):275–282. doi:10.1016/j.ceca.2003.10.010

20. Pratt JH, Ambrosius WT, Wagner MA, Maharry K. Molecular variations in the calcium-sensing receptor in relation to sodium balance and presence of hypertension in blacks and whites. Am J Hypertens. 2000;13(6 Pt 1):654-658. doi:10.1016/s0895-7061(99)00285-x

21. Muddana V, Lamb J, Greer JB, et al. Association between calcium sensing receptor gene polymorphisms and chronic pancreatitis in a US population: Role of serine protease inhibitor Kazal 1type and alcohol. World Journal of Gastroenterology. 2008;14(28):4486. doi:10.3748/wjg.14.4486

22. Lorch G, Viatchenko-Karpinski S, Ho H-T, et al. The calcium-sensing receptor is necessary for the rapid development of hypercalcemia in human lung squamous cell carcinoma. Neoplasia. 2011;13(5):428–438. doi:10.1593/neo.101620

23. Wang L, Widatalla SE, Whalen DS, Ochieng J, Sakwe AM. Association of calcium sensing receptor polymorphisms at rs1801725 with circulating calcium in breast cancer patients. BMC Cancer. 2017;17(1):511. doi:10.1186/s12885-017-3502-3

24. Schwartz GG, John EM, Rowland G, Ingles SA. Prostate cancer in African-American men and polymorphism in the calcium-sensing receptor. Cancer Biol Ther. 2010;9(12):994–999. doi:10.4161/cbt.9.12.11689

25. März W, Seelhorst U, Wellnitz B, et al. Alanine to serine polymorphism at position 986 of the calcium-sensing receptor associated with coronary heart disease, myocardial infarction, all-cause, and cardiovascular mortality. J Clin Endocrinol Metab. 2007;92(6):2363–2369. doi:10.1210/jc.2006-0071

26. Scillitani A, Guarnieri V, De Geronimo S, et al. Blood ionized calcium is associated with clustered polymorphisms in the carboxyl-terminal tail of the calcium-sensing receptor. J Clin Endocrinol Metab. 2004;89(11):5634–5638. doi:10.1210/jc.2004-0129

27. Cole DE, Vieth R, Trang HM, Wong BY, Hendy GN, Rubin LA. Association between total serum calcium and the A986S polymorphism of the calcium-sensing receptor gene. Mol Genet Metab. 2001;72(2):168–174. doi:10.1006/mgme.2000.3126

28. Vezzoli G, Scillitani A, Corbetta S, et al. Risk of nephrolithiasis in primary hyperparathyroidism is associated with two polymorphisms of the calcium-sensing receptor gene. J Nephrol. 2015;28(1):67–72. doi:10.1007/s40620-014-0106-8

29. Roden DM, Pulley JM, Basford MA, et al. Development of a large-scale de-identified DNA biobank to enable personalized medicine. Clin Pharmacol Ther. 2008;84(3):362–369. doi:10.1038/clpt.2008.89

30. Denny JC, Bastarache L, Ritchie MD, et al. Systematic comparison of phenome-wide association study of electronic medical record data and genome-wide association study data. Nat Biotechnol. 2013;31(12):1102–1110. doi:10.1038/nbt.2749

31. Dennis JK, Sealock JM, Straub P, et al. Clinical laboratory test-wide association scan of polygenic scores identifies biomarkers of complex disease. Genome Med. 2021;13(1):6. doi:10.1186/s13073-020-00820-8

32. Bien SA, Wojcik GL, Zubair N, et al. Strategies for Enriching Variant Coverage in Candidate Disease Loci on a Multiethnic Genotyping Array. PLoS One. 2016;11(12):e0167758. doi:10.1371/journal.pone.0167758

33. Abraham G, Qiu Y, Inouye M. FlashPCA2: principal component analysis of Biobank-scale genotype datasets. Bioinformatics. 2017;33(17):2776–2778. doi:10.1093/bioinformatics/btx299

34. Saidak Z, Boudot C, Abdoune R, et al. Extracellular calcium promotes the migration of breast cancer cells through the activation of the calcium sensing receptor. Exp Cell Res. 2009;315(12):2072–2080. doi:10.1016/j.yexcr.2009.03.003

35. Kumar R, Tebben PJ, Thompson JR. Vitamin D and the kidney. Arch Biochem Biophys. 2012;523(1):77–86. doi:10.1016/j.abb.2012.03.003

36. Silva BC, Bilezikian JP. Parathyroid hormone: anabolic and catabolic actions on the skeleton. Curr Opin Pharmacol. 2015;22:41–50. doi:10.1016/j.coph.2015.03.005

37. Huang H, Li T, Liao D, Zhu Z, Dong Y. Quantitative assessment of the clinical susceptibility of calcium-sensing receptor polymorphisms in cancer patients. Cancer Manag Res. 2018;10:755–763. doi:10.2147/CMAR.S147751

38. Kapur K, Johnson T, Beckmann ND, et al. Genome-wide meta-analysis for serum calcium identifies significantly associated SNPs near the calcium-sensing receptor (CASR) gene. PLoS Genet. 2010;6(7):e1001035. doi:10.1371/journal.pgen.1001035

39. Cole DEC, Vieth R, Trang HM, Wong BY-L, Hendy GN, Rubin LA. Association between Total Serum Calcium and the A986S Polymorphism of the Calcium-Sensing Receptor Gene. Molecular Genetics and Metabolism. 2001;72(2):168–174. doi:10.1006/mgme.2000.3126

40. Zagzag J, Hu MI, Fisher SB, Perrier ND. Hypercalcemia and cancer: Differential diagnosis and treatment. CA Cancer J Clin. 2018;68(5):377–386. doi:10.3322/caac.21489

