## Supplementary Material for "Calcium-Sensing Receptor Polymorphisms at rs1801725 are Associated with Increased Risk of Secondary Malignancies"

This document contains supplementary material for:

**Supplementary Methods and Results**

*Power analysis*

A power analysis demonstrated that the African dataset, though smaller, remained well powered (100%) to detect the same effect sizes observed in the larger European descent sample.  A standard normal distribution was assumed in the z-test family logistic regression function of G*Power 3.1.9.6. All regression analyses were performed using R 3.6.0.

*Phenome-wide association study (PheWAS)*

We performed a PheWAS of rs1801725 and rs1801726 as an additional secondary analysis to examine the effects of these two *CASR* SNPs across 1,858 phenotypes in the VUMC medical phenome (**Figures S1-4**). The additive and recessive models were fitted to the European and African descent datasets. Covariates included median age, sex, and the top 10 principal components for each genotyped cohort. All analyses were completed using the PheWAS 0.99.5-2 package in R 3.6.0.

*Disease phenotypes associated with polymorphisms at CASR rs1801725 and rs1801726.*

We performed an exploratory analysis on polymorphic variants of *CASR* at rs1801725 and rs1801726 in a PheWAS model to test if these variants were associated with calcium related disease phenotypes across the medical phenome. We show that several circulating calcium associated cancer phenotypes passed a p-value threshold of 0.05 in the additive PheWAS model (**Figure S2a**). These include secondary malignancy of respiratory organs (OR = 1.21, 95% CI = 1.08-1.35, p = 9.32e-04), secondary malignancy of bone (OR = 1.20, 95% CI = 1.06-1.36, p = 4.24e-03), and hypercalcemia (OR = 1.22, 95% C I = 1.06-1.40, p = 5.53e=03). Using a recessive PheWAS model, we show that polymorphisms at rs1801725 were significantly (p < 3.64E-5) associated with deficiency of humoral immunity (OR = 1.21, 95% CI = 1.08-1.35, p = 9.32e-04) in the European dataset (**Figures S1a**). We also demonstrate that polymorphisms at this locus are nominally associated (p < 0.05) with bone-related cancers (secondary malignancy of bone and cancer of bone and connective tissues) and lymphomas (non-Hodgkin’s, acute lymphoid and chronic lymphoid leukemias).

Phenome-wide significant effects were not observed in the African dataset for either of the CaSR SNPs nor were there any nominal associations with cancer phenotypes. However, we observed a modest association between rs1801275 and osteoporosis (OR = 1.41, p = 0.02) (**Figure S3-4**).

**Supplementary Table 1. Description of cancer phecodes used in this study.**

| Primary Cancer Phecodes | |
| --- | --- |
| **Phecode** | **Phenotype** |
| 145 | Cancer of mouth |
| 149 | Cancer of larynx, pharynx, nasal cavities |
| 150 | Cancer of esophagus |
| 151 | Cancer of stomach |
| 153 | Colorectal cancer |
| 155 | Cancer of liver and intrahepatic bile duct |
| 157 | Pancreatic cancer |
| 164 | Cancer of intrathoracic organs |
| 165 | Cancer within the respiratory system |
| 170 | Cancer of bone and connective tissue |
| 172 | Skin cancer |
| 174 | Breast cancer |
| 180 | Cervical cancer and dysplasia |
| 184 | Cancer of other female genital organs |
| 185 | Cancer of prostate |
| 187 | Cancer of other male genital organs |
| 189 | Cancer of urinary organs (incl. kidney and bladder) |
| 190 | Cancer of eye |
| 193 | Thyroid cancer |
| 194 | Cancer of other endocrine glands |
| 195 | Cancer suspected or other |
| 202 | Cancer of other lymphoid histiocytic tissue |
| **Secondary Malignancy Phecodes** | |
| **Phecode** | **Phenotype** |
| 198.1 | Secondary malignancy of lymph nodes |
| 198.2 | Secondary malignancy of respiratory organs |
| 198.3 | Secondary malignant neoplasm of digestive systems |
| 198.4 | Secondary malignant neoplasm of liver |
| 198.5 | Secondary malignancy of brain/spine |
| 198.6 | Secondary malignancy of bone |
| 198.7 | Secondary malignant neoplasm of skin |

| Phenotype | Estimate | SE | Z Value | P | LCI | OR | UCI | Controls | Cases | N |
| --- | --- | --- | --- | --- | --- | --- | --- | --- | --- | --- |
| Cancer of mouth | -0.27 | 0.02 | -11.09 | 1.33E-28 | 0.73 | 0.77 | 0.80 | 459696 | 2089 | 461785 |
| Cancer of larynx | -0.27 | 0.02 | -13.88 | 8.45E-44 | 0.73 | 0.76 | 0.79 | 459696 | 3170 | 462866 |
| Cancer of esophagus | -0.62 | 0.04 | -14.22 | 6.57E-46 | 0.49 | 0.54 | 0.59 | 441785 | 608 | 442393 |
| Cancer of stomach | -0.60 | 0.04 | -15.12 | 1.13E-51 | 0.51 | 0.55 | 0.59 | 441785 | 751 | 442536 |
| Colorectal cancer | -0.20 | 0.02 | -12.58 | 2.88E-36 | 0.79 | 0.82 | 0.84 | 420360 | 4677 | 425037 |
| Cancer of liver and intrahepatic bile duct | -0.57 | 0.03 | -20.30 | 1.30E-91 | 0.54 | 0.57 | 0.60 | 441785 | 1501 | 443286 |
| Pancreatic cancer | -0.61 | 0.03 | -18.30 | 8.55E-75 | 0.51 | 0.55 | 0.58 | 441785 | 1078 | 442863 |
| Cancer of intrathoracic organs | -0.21 | 0.07 | -3.17 | 1.51E-03 | 0.71 | 0.81 | 0.92 | 459911 | 281 | 460192 |
| Cancer within the respiratory system | -0.18 | 0.02 | -11.12 | 9.83E-29 | 0.81 | 0.83 | 0.86 | 459911 | 4594 | 464505 |
| Cancer of bone and connective tissue | -0.36 | 0.02 | -18.33 | 5.18E-75 | 0.67 | 0.70 | 0.72 | 462114 | 3098 | 465212 |
| Skin cancer | 0.21 | 0.01 | 24.83 | 4.79E-136 | 1.21 | 1.23 | 1.25 | 418630 | 18368 | 436998 |
| Breast cancer | 0.23 | 0.01 | 20.25 | 3.84E-91 | 1.23 | 1.26 | 1.29 | 432011 | 9672 | 441683 |
| Cervical cancer and dysplasia | -0.17 | 0.02 | -10.27 | 9.28E-25 | 0.82 | 0.84 | 0.87 | 423209 | 4496 | 427705 |
| Cancer of other female genital organs | -0.13 | 0.03 | -4.94 | 7.99E-07 | 0.84 | 0.88 | 0.93 | 439272 | 1914 | 441186 |
| Cancer of prostate | 0.17 | 0.01 | 13.33 | 1.58E-40 | 1.15 | 1.18 | 1.21 | 431562 | 8155 | 439717 |
| Cancer of other male genital organs | 0.07 | 0.04 | 1.72 | 0.09 | 0.99 | 1.07 | 1.16 | 455311 | 747 | 456058 |
| Cancer of urinary organs (incl. kidney and bladder) | -0.03 | 0.01 | -2.36 | 0.02 | 0.94 | 0.97 | 0.99 | 459109 | 5906 | 465015 |
| Cancer of eye | 0.04 | 0.05 | 0.71 | 0.48 | 0.94 | 1.04 | 1.15 | 457010 | 425 | 457435 |
| Thyroid cancer | -0.17 | 0.02 | -8.52 | 1.60E-17 | 0.81 | 0.84 | 0.87 | 456165 | 2971 | 459136 |
| Cancer of other endocrine glands | -0.26 | 0.05 | -5.00 | 5.88E-07 | 0.70 | 0.77 | 0.85 | 456165 | 452 | 456617 |
| Cancer suspected or other | -0.28 | 0.01 | -23.18 | 7.06E-119 | 0.74 | 0.75 | 0.77 | 400880 | 8270 | 409150 |
| Cancer of other lymphoid | -0.18 | 0.02 | -11.26 | 2.12E-29 | 0.81 | 0.84 | 0.86 | 450076 | 4921 | 454997 |

**Supplementary Table 2. Relationship between circulating total calcium and primary cancer diagnosis at various pathological sites.**

Logistic regression models were performed on cancer phenotypes and age adjusted inverse normalized calcium levels. The models were adjusted for sex and race. Significant results that passed Bonferroni correction (p < 3.38e-04) are highlighted. LCI = lower confidence interval; OR = odds ratio; UCI = upper confidence interval.

**Supplementary Table 3. Relationship between circulating ionized calcium and primary cancer diagnosis at various pathological sites.**

| Phenotype | Estimate | SE | Z Value | P | LCI | OR | UCI | Controls | Cases | N |
| --- | --- | --- | --- | --- | --- | --- | --- | --- | --- | --- |
| Cancer of mouth | -0.12 | 0.03 | -3.54 | 3.98E-04 | 0.83 | 0.89 | 0.95 | 81125 | 1020 | 82145 |
| Cancer of larynx | -0.13 | 0.03 | -4.64 | 3.57E-06 | 0.83 | 0.88 | 0.93 | 81125 | 1415 | 82540 |
| Cancer of esophagus | -0.10 | 0.06 | -1.80 | 0.07 | 0.81 | 0.90 | 1.01 | 74622 | 358 | 74980 |
| Cancer of stomach | -0.11 | 0.05 | -2.21 | 0.03 | 0.81 | 0.89 | 0.99 | 74622 | 414 | 75036 |
| Colorectal cancer | -0.18 | 0.03 | -7.09 | 1.38E-12 | 0.80 | 0.84 | 0.88 | 72383 | 1731 | 74114 |
| Cancer of liver and intrahepatic bile duct | 0.18 | 0.04 | 4.80 | 1.55E-06 | 1.11 | 1.19 | 1.28 | 74622 | 828 | 75450 |
| Pancreatic cancer | -0.23 | 0.04 | -5.36 | 8.33E-08 | 0.73 | 0.79 | 0.86 | 74622 | 576 | 75198 |
| Cancer of intrathoracic organs | 0.02 | 0.09 | 0.19 | 0.85 | 0.86 | 1.02 | 1.20 | 80752 | 148 | 80900 |
| Cancer within the respiratory system | 0.01 | 0.02 | 0.25 | 0.80 | 0.96 | 1.01 | 1.05 | 80752 | 1955 | 82707 |
| Cancer of bone and connective tissue | -0.21 | 0.03 | -7.70 | 1.35E-14 | 0.77 | 0.81 | 0.86 | 81401 | 1515 | 82916 |
| Skin cancer | 0.07 | 0.02 | 4.33 | 1.47E-05 | 1.04 | 1.07 | 1.11 | 74506 | 4215 | 78721 |
| Breast cancer | 0.02 | 0.02 | 0.75 | 0.45 | 0.97 | 1.02 | 1.07 | 77908 | 1765 | 79673 |
| Cervical cancer and dysplasia | -0.03 | 0.04 | -0.61 | 0.54 | 0.90 | 0.98 | 1.06 | 77164 | 612 | 77776 |
| Cancer of other female genital organs | -0.29 | 0.04 | -7.16 | 8.16E-13 | 0.69 | 0.74 | 0.81 | 78633 | 612 | 79245 |
| Cancer of prostate | -0.19 | 0.03 | -7.70 | 1.31E-14 | 0.78 | 0.82 | 0.87 | 74796 | 1919 | 76715 |
| Cancer of other male genital organs | -0.13 | 0.08 | -1.63 | 0.10 | 0.76 | 0.88 | 1.03 | 80719 | 195 | 80914 |
| Cancer of urinary organs (incl. kidney and bladder) | -0.29 | 0.02 | -14.11 | 3.17E-45 | 0.72 | 0.75 | 0.78 | 80201 | 2833 | 83034 |
| Cancer of eye | -0.12 | 0.09 | -1.27 | 0.20 | 0.74 | 0.89 | 1.07 | 79423 | 121 | 79544 |
| Thyroid cancer | -0.20 | 0.03 | -7.09 | 1.30E-12 | 0.78 | 0.82 | 0.87 | 78983 | 1434 | 80417 |
| Cancer of other endocrine glands | -0.33 | 0.06 | -5.29 | 1.23E-07 | 0.64 | 0.72 | 0.81 | 78983 | 280 | 79263 |
| Cancer suspected or other | -0.11 | 0.02 | -6.44 | 1.22E-10 | 0.87 | 0.90 | 0.93 | 62274 | 3926 | 66200 |
| Cancer of other lymphoid | 0.03 | 0.03 | 1.25 | 0.21 | 0.98 | 1.03 | 1.09 | 77518 | 1655 | 79173 |

Logistic regression models were performed on primary cancer phenotypes and age adjusted inverse normalized ionized calcium levels. The models were adjusted for sex and race. Significant results that passed Bonferroni correction (p < 3.38e-04) are highlighted. LCI = lower confidence interval; OR = odds ratio; UCI = upper confidence interval.

**Supplementary Table 4. Relationship between circulating vitamin D and primary cancer diagnosis at various pathological sites.**

| Phenotype | Estimate | SE | Z Value | P | LCI | OR | UCI | Controls | Cases | N |
| --- | --- | --- | --- | --- | --- | --- | --- | --- | --- | --- |
| Cancer of mouth | -0.02 | 0.06 | -0.35 | 0.73 | 0.87 | 0.98 | 1.10 | 103713 | 336 | 104049 |
| Cancer of larynx | -0.01 | 0.05 | -0.13 | 0.90 | 0.90 | 0.99 | 1.09 | 103713 | 485 | 104198 |
| Cancer of esophagus | -0.36 | 0.10 | -3.66 | 2.49E-04 | 0.58 | 0.70 | 0.85 | 97392 | 124 | 97516 |
| Cancer of stomach | -0.38 | 0.08 | -4.94 | 7.95E-07 | 0.59 | 0.68 | 0.80 | 97392 | 196 | 97588 |
| Colorectal cancer | -0.13 | 0.03 | -4.65 | 3.26E-06 | 0.83 | 0.88 | 0.93 | 85915 | 1517 | 87432 |
| Cancer of liver and intrahepatic bile duct | -0.14 | 0.05 | -3.05 | 2.26E-03 | 0.79 | 0.87 | 0.95 | 97392 | 551 | 97943 |
| Pancreatic cancer | -0.37 | 0.07 | -5.43 | 5.78E-08 | 0.61 | 0.69 | 0.79 | 97392 | 253 | 97645 |
| Cancer of intrathoracic organs | -0.09 | 0.13 | -0.73 | 0.47 | 0.71 | 0.91 | 1.17 | 103987 | 67 | 104054 |
| Cancer within the respiratory system | -0.26 | 0.04 | -7.15 | 8.55E-13 | 0.72 | 0.77 | 0.83 | 103987 | 867 | 104854 |
| Cancer of bone and connective tissue | -0.08 | 0.05 | -1.67 | 0.09 | 0.84 | 0.92 | 1.01 | 104414 | 513 | 104927 |
| Skin cancer | 0.06 | 0.01 | 4.25 | 2.17E-05 | 1.03 | 1.06 | 1.09 | 88268 | 6219 | 94487 |
| Breast cancer | 0.12 | 0.02 | 7.44 | 9.89E-14 | 1.10 | 1.13 | 1.17 | 92103 | 4237 | 96340 |
| Cervical cancer and dysplasia | -0.05 | 0.03 | -1.64 | 0.10 | 0.90 | 0.95 | 1.01 | 89766 | 1383 | 91149 |
| Cancer of other female genital organs | -0.05 | 0.05 | -1.03 | 0.30 | 0.87 | 0.95 | 1.04 | 95568 | 531 | 96099 |
| Cancer of prostate | 0.05 | 0.03 | 1.53 | 0.13 | 0.99 | 1.05 | 1.11 | 96306 | 1453 | 97759 |
| Cancer of other male genital organs | -0.12 | 0.10 | -1.27 | 0.20 | 0.74 | 0.89 | 1.07 | 103108 | 135 | 103243 |
| Cancer of urinary organs (incl. kidney and bladder) | -0.24 | 0.03 | -7.77 | 7.75E-15 | 0.74 | 0.79 | 0.84 | 103536 | 1223 | 104759 |
| Cancer of eye | -0.41 | 0.12 | -3.26 | 1.10E-03 | 0.52 | 0.67 | 0.85 | 102748 | 72 | 102820 |
| Thyroid cancer | 0.16 | 0.03 | 5.21 | 1.92E-07 | 1.10 | 1.17 | 1.25 | 101088 | 1199 | 102287 |
| Cancer of other endocrine glands | -0.06 | 0.09 | -0.64 | 0.52 | 0.78 | 0.94 | 1.13 | 101088 | 128 | 101216 |
| Cancer suspected or other | -0.20 | 0.02 | -8.41 | 4.18E-17 | 0.79 | 0.82 | 0.86 | 84362 | 2185 | 86547 |
| Cancer of other lymphoid | -0.11 | 0.03 | -4.03 | 5.56E-05 | 0.84 | 0.89 | 0.94 | 99708 | 1452 | 101160 |

Logistic regression models were performed on primary cancers and age adjusted inverse normalized vitamin D levels. The models were adjusted for sex and race. Significant results that passed Bonferroni correction (p < 3.38e-04) are highlighted. LCI = lower confidence interval; OR = odds ratio; UCI = upper confidence interval.

**Supplementary Table 5. Relationship between circulating intact parathyroid hormone and primary cancer diagnosis at various pathological sites.**

| Phenotype | Estimate | SE | Z Value | P | LCI | OR | UCI | Controls | Cases | N |
| --- | --- | --- | --- | --- | --- | --- | --- | --- | --- | --- |
| Cancer of mouth | -0.25 | 0.08 | -3.08 | 2.08E-03 | 0.66 | 0.78 | 0.91 | 33928 | 156 | 34084 |
| Cancer of larynx | -0.33 | 0.07 | -4.79 | 1.64E-06 | 0.63 | 0.72 | 0.82 | 33928 | 217 | 34145 |
| Cancer of esophagus | -0.14 | 0.14 | -0.99 | 0.32 | 0.66 | 0.87 | 1.14 | 31427 | 53 | 31480 |
| Cancer of stomach | 0.20 | 0.11 | 1.79 | 0.07 | 0.98 | 1.23 | 1.53 | 31427 | 83 | 31510 |
| Colorectal cancer | -0.04 | 0.05 | -0.76 | 0.45 | 0.87 | 0.96 | 1.06 | 28218 | 451 | 28669 |
| Cancer of liver and intrahepatic bile duct | -0.08 | 0.09 | -0.87 | 0.38 | 0.78 | 0.93 | 1.10 | 31427 | 132 | 31559 |
| Pancreatic cancer | 0.10 | 0.09 | 1.07 | 0.28 | 0.92 | 1.10 | 1.32 | 31427 | 131 | 31558 |
| Cancer of intrathoracic organs | -0.07 | 0.18 | -0.37 | 0.71 | 0.66 | 0.94 | 1.34 | 33946 | 33 | 33979 |
| Cancer within the respiratory system | -0.18 | 0.05 | -3.55 | 3.93E-04 | 0.76 | 0.84 | 0.92 | 33946 | 437 | 34383 |
| Cancer of bone and connective tissue | -0.26 | 0.07 | -3.93 | 8.48E-05 | 0.68 | 0.77 | 0.88 | 34225 | 251 | 34476 |
| Skin cancer | -0.06 | 0.02 | -2.63 | 8.66E-03 | 0.91 | 0.94 | 0.99 | 28611 | 2511 | 31122 |
| Breast cancer | -0.16 | 0.03 | -4.91 | 9.26E-07 | 0.80 | 0.85 | 0.91 | 30365 | 1216 | 31581 |
| Cervical cancer and dysplasia | 0.12 | 0.05 | 2.20 | 0.03 | 1.01 | 1.13 | 1.25 | 29889 | 409 | 30298 |
| Cancer of other female genital organs | -0.01 | 0.07 | -0.14 | 0.89 | 0.86 | 0.99 | 1.14 | 31420 | 219 | 31639 |
| Cancer of prostate | -0.06 | 0.04 | -1.51 | 0.13 | 0.87 | 0.94 | 1.02 | 30910 | 648 | 31558 |
| Cancer of other male genital organs | -0.01 | 0.13 | -0.07 | 0.94 | 0.77 | 0.99 | 1.27 | 33601 | 62 | 33663 |
| Cancer of urinary organs (incl. kidney and bladder) | 0.22 | 0.03 | 6.79 | 1.16E-11 | 1.17 | 1.25 | 1.33 | 33151 | 1034 | 34185 |
| Cancer of eye | 0.15 | 0.17 | 0.87 | 0.38 | 0.83 | 1.16 | 1.62 | 33678 | 39 | 33717 |
| Thyroid cancer | -0.76 | 0.03 | -22.82 | 2.76E-115 | 0.44 | 0.47 | 0.50 | 31033 | 1128 | 32161 |
| Cancer of other endocrine glands | 0.18 | 0.12 | 1.42 | 0.16 | 0.93 | 1.19 | 1.52 | 31033 | 70 | 31103 |
| Cancer suspected or other | -0.22 | 0.03 | -6.31 | 2.73E-10 | 0.75 | 0.80 | 0.86 | 25980 | 949 | 26929 |
| Cancer of other lymphoid | -0.13 | 0.04 | -3.00 | 2.71E-03 | 0.81 | 0.88 | 0.96 | 32164 | 590 | 32754 |

Logistic regression models were performed on primary cancers and age adjusted inverse normalized parathyrin intact levels. The models were adjusted for sex and race. Significant results that passed Bonferroni correction (p < 3.38e-04) are highlighted. LCI = lower confidence interval; OR = odds ratio; UCI = upper confidence interval.

**Supplementary Table 6. Association of circulating calcium, vitamin D and parathyroid hormone with secondary malignancies.**

| Phenotype | Estimate | SE | Z Value | P | LCI | OR | UCI | Phecode | Controls | Cases | N |
| --- | --- | --- | --- | --- | --- | --- | --- | --- | --- | --- | --- |
| Calcium | | | | | | | | | | | |
| Secondary malignancy of lymph nodes | -0.13 | 0.01 | -9.34 | 9.48E-21 | 0.86 | 0.88 | 0.90 | 400880 | 6716 | 407596 | -0.13 |
| Secondary malignancy of respiratory organs | -0.23 | 0.02 | -12.81 | 1.45E-37 | 0.77 | 0.79 | 0.82 | 400880 | 3759 | 404639 | -0.23 |
| Secondary malignant neoplasm of digestive systems | -0.50 | 0.03 | -19.24 | 1.63E-82 | 0.58 | 0.61 | 0.64 | 400880 | 1779 | 402659 | -0.50 |
| Secondary malignant neoplasm of liver | -0.37 | 0.02 | -18.24 | 2.72E-74 | 0.66 | 0.69 | 0.72 | 400880 | 2926 | 403806 | -0.37 |
| Secondary malignancy of brain/spine | -0.29 | 0.03 | -11.21 | 3.70E-29 | 0.71 | 0.75 | 0.79 | 400880 | 1794 | 402674 | -0.29 |
| Secondary malignancy of bone | -0.26 | 0.02 | -13.90 | 6.09E-44 | 0.75 | 0.77 | 0.80 | 400880 | 3530 | 404410 | -0.26 |
| Secondary malignant neoplasm of skin | -0.07 | 0.04 | -1.71 | 0.09 | 0.86 | 0.93 | 1.01 | 400880 | 725 | 401605 | -0.07 |
| Ionized Calcium | | | | | | | | | | | |
| Secondary malignancy of lymph nodes | -0.17 | 0.02 | -8.22 | 2.00E-16 | 0.81 | 0.85 | 0.88 | 62274 | 2740 | 65014 | -0.17 |
| Secondary malignancy of respiratory organs | -0.14 | 0.02 | -5.75 | 9.17E-09 | 0.83 | 0.87 | 0.91 | 62274 | 1764 | 64038 | -0.14 |
| Secondary malignant neoplasm of digestive systems | -0.32 | 0.03 | -9.65 | 5.06E-22 | 0.68 | 0.73 | 0.78 | 62274 | 988 | 63262 | -0.32 |
| Secondary malignant neoplasm of liver | -0.19 | 0.03 | -6.57 | 5.01E-11 | 0.78 | 0.83 | 0.88 | 62274 | 1328 | 63602 | -0.19 |
| Secondary malignancy of brain/spine | -0.20 | 0.03 | -5.82 | 5.98E-09 | 0.77 | 0.82 | 0.88 | 62274 | 947 | 63221 | -0.20 |
| Secondary malignancy of bone | -0.08 | 0.03 | -3.25 | 1.17E-03 | 0.88 | 0.92 | 0.97 | 62274 | 1656 | 63930 | -0.08 |
| Secondary malignant neoplasm of skin | -0.11 | 0.06 | -1.74 | 0.08 | 0.79 | 0.90 | 1.01 | 62274 | 267 | 62541 | -0.11 |
| Vitamin D | | | | | | | | | | | |
| Secondary malignancy of lymph nodes | -0.05 | 0.03 | -1.83 | 0.07 | 0.91 | 0.96 | 1.00 | 84362 | 1851 | 86213 | -0.05 |
| Secondary malignancy of respiratory organs | -0.29 | 0.04 | -6.89 | 5.45E-12 | 0.69 | 0.75 | 0.82 | 84362 | 675 | 85037 | -0.29 |
| Secondary malignant neoplasm of digestive systems | -0.33 | 0.06 | -5.82 | 5.83E-09 | 0.64 | 0.72 | 0.80 | 84362 | 361 | 84723 | -0.33 |
| Secondary malignant neoplasm of liver | -0.33 | 0.04 | -7.57 | 3.84E-14 | 0.66 | 0.72 | 0.78 | 84362 | 605 | 84967 | -0.33 |
| Secondary malignancy of brain/spine | -0.27 | 0.06 | -4.24 | 2.28E-05 | 0.67 | 0.76 | 0.86 | 84362 | 283 | 84645 | -0.27 |
| Secondary malignancy of bone | -0.18 | 0.04 | -4.58 | 4.66E-06 | 0.77 | 0.84 | 0.90 | 84362 | 748 | 85110 | -0.18 |
| Secondary malignant neoplasm of skin | -0.30 | 0.10 | -3.16 | 1.58E-03 | 0.61 | 0.74 | 0.89 | 84362 | 125 | 84487 | -0.30 |
| Parathyroid Hormone | | | | | | | | | | | |
| Secondary malignancy of lymph nodes | -0.33 | 0.04 | -8.61 | 7.09E-18 | 0.67 | 0.72 | 0.78 | 25980 | 790 | 26770 | -0.33 |
| Secondary malignancy of respiratory organs | -0.25 | 0.05 | -4.61 | 4.00E-06 | 0.70 | 0.78 | 0.87 | 25980 | 379 | 26359 | -0.25 |
| Secondary malignant neoplasm of digestive systems | -0.16 | 0.08 | -2.04 | 0.04 | 0.73 | 0.85 | 0.99 | 25980 | 180 | 26160 | -0.16 |
| Secondary malignant neoplasm of liver | -0.24 | 0.06 | -3.90 | 9.68E-05 | 0.70 | 0.79 | 0.89 | 25980 | 305 | 26285 | -0.24 |
| Secondary malignancy of brain/spine | -0.38 | 0.09 | -4.27 | 1.96E-05 | 0.57 | 0.68 | 0.81 | 25980 | 141 | 26121 | -0.38 |
| Secondary malignancy of bone | -0.38 | 0.05 | -7.22 | 5.26E-13 | 0.62 | 0.68 | 0.76 | 25980 | 396 | 26376 | -0.38 |
| Secondary malignant neoplasm of skin | -0.31 | 0.12 | -2.54 | 0.01 | 0.58 | 0.74 | 0.93 | 25980 | 79 | 26059 | -0.31 |

The logistic regressions models were adjusted for sex, median age, and principal components 1-10. Significant results that passed Bonferroni correction (p < 3.38e-04) are highlighted. LCI = lower confidence interval; OR = odds ratio; UCI = upper confidence interval.

**Supplementary Table 7.** **Associating of calcium, vitamin D and parathyroid hormone levels with polymorphisms at rs1801725 and rs1801726 in individuals of European descent.**

|  | rs1801725 | | | | rs1801726 | | | |
| --- | --- | --- | --- | --- | --- | --- | --- | --- |
|  | All | 0 RA | 1 RA | 2 RA | All | 0 RA | 1 RA | 2 RA |
| Calcium (mg/dL) | | | | | | | | |
| N | 51171 | 38094 | 12131 | 946 | 51171 | 47415 | 3675 | 79 |
| Median [IQR] | 9.30  [9.05, 9.60] | 9.30  [9.00, 9.60] | 9.40  [9.10, 9.60] | 9.50  [9.20, 9.70] | 9.30  [9.05, 9.60] | 9.30  [9.05, 9.60] | 9.30  [9.00, 9.60] | 9.40  [9.03, 9.70] |
| Ionized Calcium (mg/dL) | | | | | | | | |
| N | 13897 | 10369 | 3281 | 247 | 13897 | 12866 | 1008 | 22 |
| Median [IQR] | 4.52  [4.32, 4.73] | 4.51  [4.31, 4.73] | 4.55  [4.35, 4.77] | 4.59  [4.34, 4.79] | 4.52  [4.32, 4.73] | 4.52  [4.32, 4.73] | 4.53  [4.33, 4.74] | 4.52  [4.43, 4.69] |
| Vitamin D (ng/mL) | | | | | | | | |
| N | 18883 | 14076 | 4452 | 355 | 18883 | 17497 | 1354 | 31 |
| Median [IQR] | 31.00  [24.00, 39.00] | 31.00  [24.00, 39.00] | 31.00  [24.00, 39.00] | 32.00  [24.00, 39.75] | 31.00  [24.00, 39.00] | 31.00  [24.00, 39.00] | 31.00  [24.00, 39.00] | 28.00  [21.50, 43.25] |
| Parathyroid Hormone (pg/mL) | | | | | | | | |
| N | 6936 | 5133 | 1659 | 144 | 6936 | 6430 | 497 | 9 |
| Median [IQR] | 56.00  [36.00, 91.00] | 56.00  [36.00, 91.00] | 56.00  [35.00, 88.25] | 59.50  [39.75, 106.25] | 56.00  [36.00, 91.00] | 56.00  [36.00, 91.00] | 55.50  [36.00, 88.50] | 46.00  [39.00, 205.00] |

Median (a) calcium, (b) ionized calcium, (c) vitamin D, and (d) parathyroid hormone levels. Wilcoxon rank sum test was used to determine statistical significance between allele carriers (p < 0.05). RA = risk alleles, IQR = interquartile range.

**Supplementary Table 8. Associating of calcium, vitamin D and parathyroid hormone levels with polymorphisms at rs1801725 and rs1801726 in individuals of African descent.**

|  | rs1801725 | | | | rs1801726 | | | |
| --- | --- | --- | --- | --- | --- | --- | --- | --- |
|  | All | 0 RA | 1 RA | 2 RA | All | 0 RA | 1 RA | 2 RA |
| Calcium (mg/dL) | | | | | | | | |
| N | 9878 | 9159 | 694 | 18 | 9878 | 6837 | 2749 | 283 |
| Median  [IQR] | 9.40  [9.10, 9.60] | 9.40  [9.10, 9.60] | 9.40  [9.20, 9.70] | 9.43  [9.22, 9.88] | 9.40  [9.10, 9.60] | 9.40  [9.10, 9.60] | 9.40  [9.10, 9.60] | 9.40  [9.10, 9.60] |
| Ionized Calcium (mg/dL) | | | | | | | | |
| N | 2336 | 2175 | 156 | 4 | 2336 | 1561 | 709 | 66 |
| Median  [IQR] | 4.57  [4.35, 4.80] | 4.57  [4.34, 4.80] | 4.65  [4.45, 4.89] | 4.53  [4.41, 4.79] | 4.57  [4.35, 4.80] | 4.57  [4.35, 4.80] | 4.57  [4.33, 4.81] | 4.55  [4.33, 4.77] |
| Vitamin D (ng/mL) | | | | | | | | |
| N | 3341 | 3115 | 219 | 4 | 3341 | 2289 | 924 | 124 |
| Median  [IQR] | 23.00  [17.00, 30.50] | 23.00  [16.50, 30.50] | 24.00  [18.00, 31.00] | 19.50  [18.25, 22.25] | 23.00  [17.00, 30.50] | 22.50  [16.50, 30.00] | 23.00  [16.88, 31.00] | 24.75  [18.90, 31.25] |
| Parathyroid Hormone (pg/mL) | | | | | | | | |
| N | 1430 | 1343 | 86 | 1 | 1430 | 976 | 408 | 45 |
| Median  [IQR] | 82.00  [50.0, 160.0] | 82.00  [49.0, 160.3] | 87.25  [59.6, 151.4] | 49.00  [49.00, 49.00] | 82.00  [50.00, 160.00] | 82.75  [50.4, 159.25] | 80.50  [48.00, 158.1] | 89.00  [52.00, 167.00] |

Median (a) calcium, (b) ionized calcium, (c) vitamin D, and (d) parathyroid hormone levels. Wilcoxon rank sum test was used to determine statistical significance between allele carriers (p < 0.05). RA = risk alleles, IQR = interquartile range.

**Supplementary Table 9.** **Relationship between CaSR SNPs and circulating calcium, vitamin D and parathyroid hormone levels.**

| Lab | Estimate | SE | T Value | P | LCI | OR | UCI | 0 RA | 1 RA | 2 RA |
| --- | --- | --- | --- | --- | --- | --- | --- | --- | --- | --- |
| European Genotyped Sample: rs1801725 | | | | | | | | | | |
| Calcium | 0.14 | 0.01 | 18.08 | 8.40E-73 | 1.13 | 1.15 | 1.16 | 38098 | 12132 | 946 |
| Ionized Calcium | 0.10 | 0.02 | 6.06 | 1.43E-09 | 1.07 | 1.10 | 1.14 | 10370 | 3282 | 247 |
| Vitamin D | -0.01 | 0.01 | -0.67 | 0.50 | 0.96 | 0.99 | 1.02 | 14078 | 4452 | 355 |
| Parathyroid Hormone | 0.001 | 0.02 | 0.03 | 0.98 | 0.95 | 1.00 | 1.05 | 5134 | 1659 | 144 |
| European Genotyped Sample: rs1801726 | | | | | | | | | | |
| Calcium | -0.02 | 0.01 | -1.41 | 0.16 | 0.96 | 0.98 | 1.01 | 47420 | 3675 | 79 |
| Ionized Calcium | 0.03 | 0.03 | 1.07 | 0.28 | 0.97 | 1.03 | 1.09 | 12868 | 1008 | 22 |
| Vitamin D | -0.02 | 0.03 | -0.66 | 0.51 | 0.94 | 0.98 | 1.03 | 17499 | 1354 | 31 |
| Parathyroid Hormone | 0.03 | 0.04 | 0.57 | 0.57 | 0.94 | 1.03 | 1.12 | 6431 | 497 | 9 |
| African Descent Genotyped Sample: rs1801725 | | | | | | | | | | |
| Calcium | 0.12 | 0.03 | 3.68 | 2.34E-04 | 1.06 | 1.12 | 1.20 | 9159 | 694 | 18 |
| Ionized Calcium | 0.16 | 0.08 | 1.98 | 4.73E-02 | 1.00 | 1.17 | 1.37 | 2175 | 156 | 4 |
| Vitamin D | 0.07 | 0.07 | 1.08 | 0.28 | 0.94 | 1.07 | 1.22 | 3115 | 219 | 4 |
| Parathyroid Hormone | 0.10 | 0.11 | 0.94 | 0.35 | 0.90 | 1.11 | 1.37 | 1343 | 86 | 1 |
| African Descent Genotyped Sample: rs1801726 | | | | | | | | | | |
| Calcium | 0.002 | 0.02 | 0.13 | 0.90 | 0.97 | 1.00 | 1.03 | 6837 | 2749 | 283 |
| Ionized Calcium | 0.01 | 0.04 | 0.36 | 0.72 | 0.94 | 1.01 | 1.09 | 1561 | 709 | 66 |
| Vitamin D | 0.06 | 0.03 | 1.83 | 0.07 | 1.00 | 1.06 | 1.12 | 2289 | 924 | 124 |
| Parathyroid Hormone | 0.03 | 0.05 | 0.64 | 0.52 | 0.94 | 1.03 | 1.13 | 976 | 408 | 45 |

Laboratory values were inverse normalized and adjusted for age. The regression models were adjusted for principal components 1-10. Significant results that passed Bonferroni correction (p < 3.38e-04) are highlighted. LCI = lower confidence interval; OR = odds ratio; UCI = upper confidence interval; RA = risk alleles.

**Supplementary Table 10. Relationship between circulating calcium, vitamin D and parathyroid hormone levels in CaSR rs1801725 allele carriers before and after cancer diagnosis.**

| Lab | Estimate | SE | T Value | P | LCI | OR | UCI | 0 RA | 1 RA | 2 RA |
| --- | --- | --- | --- | --- | --- | --- | --- | --- | --- | --- |
| European Genotyped Sample – Before Cancer Diagnosis Labs  rs1801725 | | | | | | | | | | |
| Calcium | 0.13 | 0.01 | 17.79 | 1.53E-70 | 1.13 | 1.14 | 1.16 | 36807 | 11746 | 917 |
| Ionized Calcium | 0.09 | 0.02 | 5.24 | 1.66E-07 | 1.06 | 1.09 | 1.13 | 8948 | 2813 | 219 |
| Vitamin D | -0.01 | 0.01 | -0.86 | 0.39 | 0.96 | 0.99 | 1.02 | 12937 | 4106 | 330 |
| Parathyroid Hormone | -0.02 | 0.02 | -0.64 | 0.52 | 0.94 | 0.98 | 1.03 | 4605 | 1495 | 126 |
| European Genotyped Sample – After Cancer Diagnosis Labs  rs1801725 | | | | | | | | | | |
| Calcium | 0.12 | 0.02 | 5.31 | 1.16E-07 | 1.08 | 1.12 | 1.17 | 5351 | 1688 | 126 |
| Ionized Calcium | 0.09 | 0.02 | 5.24 | 1.66E-07 | 1.06 | 1.09 | 1.13 | 8948 | 2813 | 219 |
| Vitamin D | 0.02 | 0.04 | 0.58 | 0.56 | 0.94 | 1.02 | 1.11 | 1707 | 547 | 37 |
| Parathyroid Hormone | 0.03 | 0.06 | 0.50 | 0.62 | 0.92 | 1.03 | 1.16 | 694 | 234 | 20 |

Logistic regressions models were performed on CaSR rs1801725 and stratified laboratory values based on the first diagnosis of a primary cancer for European descent individuals. Laboratory values were inverse normalized and adjusted for age and the regression models were adjusted for principal components 1-10. Significant results that passed Bonferroni correction (p < 3.38e-04) highlighted. LCI = lower confidence interval; OR = odds ratio; UCI = upper confidence interval; RA = risk alleles.

**Supplementary Table 11.** **Association of CaSR SNPs with secondary malignancies in European descent individuals – Additive regression analysis.**

| Phenotype | Estimate | SE | Z Value | P | LCI | OR | UCI | Controls | Cases | 0 Alleles | 1 Allele | 2 Alleles |
| --- | --- | --- | --- | --- | --- | --- | --- | --- | --- | --- | --- | --- |
| rs1801725 | | | | | | | | | | | | |
| Secondary malignancy of lymph nodes | 0.08 | 0.05 | 1.81 | 0.07 | 0.99 | 1.09 | 1.19 | 40476 | 2179 | 31719 | 10150 | 786 |
| Secondary malignancy of respiratory organs | 0.19 | 0.06 | 3.30 | 9.61E-04 | 1.08 | 1.21 | 1.35 | 40476 | 1249 | 31003 | 9958 | 764 |
| Secondary malignant neoplasm of digestive systems | 0.07 | 0.08 | 0.79 | 0.43 | 0.91 | 1.07 | 1.26 | 40476 | 615 | 30570 | 9774 | 747 |
| Secondary malignant neoplasm of liver | 0.11 | 0.07 | 1.73 | 0.08 | 0.99 | 1.12 | 1.28 | 40476 | 955 | 30808 | 9869 | 754 |
| Secondary malignancy of brain/spine | -0.01 | 0.09 | -0.13 | 0.89 | 0.83 | 0.99 | 1.18 | 40476 | 547 | 30530 | 9749 | 744 |
| Secondary malignancy of bone | 0.18 | 0.06 | 2.86 | 4.28E-03 | 1.06 | 1.20 | 1.36 | 40476 | 958 | 30806 | 9863 | 765 |
| Secondary malignant neoplasm of skin | 0.23 | 0.12 | 1.91 | 0.06 | 0.99 | 1.26 | 1.60 | 40476 | 259 | 30301 | 9693 | 741 |
| rs1801726 | | | | | | | | | | | | |
| Secondary malignancy of lymph nodes | 0.02 | 0.08 | 0.27 | 0.79 | 0.87 | 1.02 | 1.20 | 40476 | 2179 | 39517 | 3068 | 70 |
| Secondary malignancy of respiratory organs | 0.06 | 0.10 | 0.60 | 0.55 | 0.87 | 1.07 | 1.31 | 40476 | 1249 | 38655 | 3001 | 69 |
| Secondary malignant neoplasm of digestive systems | -0.01 | 0.15 | -0.07 | 0.94 | 0.73 | 0.99 | 1.33 | 40476 | 615 | 38072 | 2953 | 66 |
| Secondary malignant neoplasm of liver | -0.13 | 0.13 | -1.01 | 0.31 | 0.68 | 0.88 | 1.13 | 40476 | 955 | 38396 | 2967 | 68 |
| Secondary malignancy of brain/spine | -0.08 | 0.17 | -0.50 | 0.62 | 0.66 | 0.92 | 1.27 | 40476 | 547 | 38014 | 2941 | 68 |
| Secondary malignancy of bone | -0.09 | 0.13 | -0.69 | 0.49 | 0.71 | 0.92 | 1.17 | 40476 | 958 | 38397 | 2968 | 69 |
| Secondary malignant neoplasm of skin | 0.16 | 0.21 | 0.73 | 0.47 | 0.77 | 1.17 | 1.78 | 40476 | 259 | 37740 | 2928 | 67 |

The models were adjusted for median age, sex, and principal components 1-10. Significant results that passed a significance threshold of 0.05 are highlighted. LCI = lower confidence interval; OR = odds ratio; UCI = upper confidence interval.

**Supplementary Table 12. Association of CaSR SNPs with secondary malignancies in European descent individuals – Recessive regression analysis**

| Phenotype | Estimate | SE | Z Value | P | LCI | OR | UCI | Controls | Cases | 0 Alleles | 1 Allele |
| --- | --- | --- | --- | --- | --- | --- | --- | --- | --- | --- | --- |
| rs1801725 | | | | | | | | | | | |
| Secondary malignancy of lymph nodes | 0.30 | 0.15 | 2.04 | 0.04 | 1.01 | 1.36 | 1.82 | 40476 | 2179 | 41869 | 786 |
| Secondary malignancy of respiratory organs | 0.29 | 0.19 | 1.49 | 0.14 | 0.91 | 1.33 | 1.95 | 40476 | 1249 | 40961 | 764 |
| Secondary malignant neoplasm of digestive systems | 0.10 | 0.30 | 0.33 | 0.74 | 0.62 | 1.10 | 1.97 | 40476 | 615 | 40344 | 747 |
| Secondary malignant neoplasm of liver | 0.12 | 0.24 | 0.52 | 0.61 | 0.71 | 1.13 | 1.80 | 40476 | 955 | 40677 | 754 |
| Secondary malignancy of brain/spine | -0.05 | 0.34 | -0.15 | 0.88 | 0.49 | 0.95 | 1.85 | 40476 | 547 | 40279 | 744 |
| Secondary malignancy of bone | 0.60 | 0.19 | 3.12 | 1.80E-03 | 1.25 | 1.82 | 2.65 | 40476 | 958 | 40669 | 765 |
| Secondary malignant neoplasm of skin | 0.32 | 0.42 | 0.77 | 0.44 | 0.61 | 1.38 | 3.13 | 40476 | 259 | 39994 | 741 |
| rs1801726 | | | | | | | | | | | |
| Secondary malignancy of lymph nodes | 0.21 | 0.52 | 0.41 | 0.68 | 0.44 | 1.24 | 3.44 | 40476 | 2179 | 42585 | 70 |
| Secondary malignancy of respiratory organs | 0.53 | 0.60 | 0.89 | 0.37 | 0.53 | 1.70 | 5.50 | 40476 | 1249 | 41656 | 69 |
| Secondary malignant neoplasm of digestive systems | -11.23 | 174.62 | -0.06 | 0.95 | 3.02E-154 | 1.33E-05 | 5.84E+143 | 40476 | 615 | 41025 | 66 |
| Secondary malignant neoplasm of liver | 0.37 | 0.72 | 0.51 | 0.61 | 0.35 | 1.45 | 5.98 | 40476 | 955 | 41363 | 68 |
| Secondary malignancy of brain/spine | 0.92 | 0.72 | 1.27 | 0.20 | 0.61 | 2.51 | 10.36 | 40476 | 547 | 40955 | 68 |
| Secondary malignancy of bone | 0.79 | 0.60 | 1.32 | 0.19 | 0.68 | 2.20 | 7.11 | 40476 | 958 | 41365 | 69 |
| Secondary malignant neoplasm of skin | 0.98 | 1.02 | 0.96 | 0.34 | 0.36 | 2.65 | 19.41 | 40476 | 259 | 40668 | 67 |

The models were adjusted for median age, sex, and principal components 1-10. Significant results that passed Bonferroni correction (p < 3.38e-04) are highlighted. LCI = lower confidence interval; OR = odds ratio; UCI = upper confidence interval.

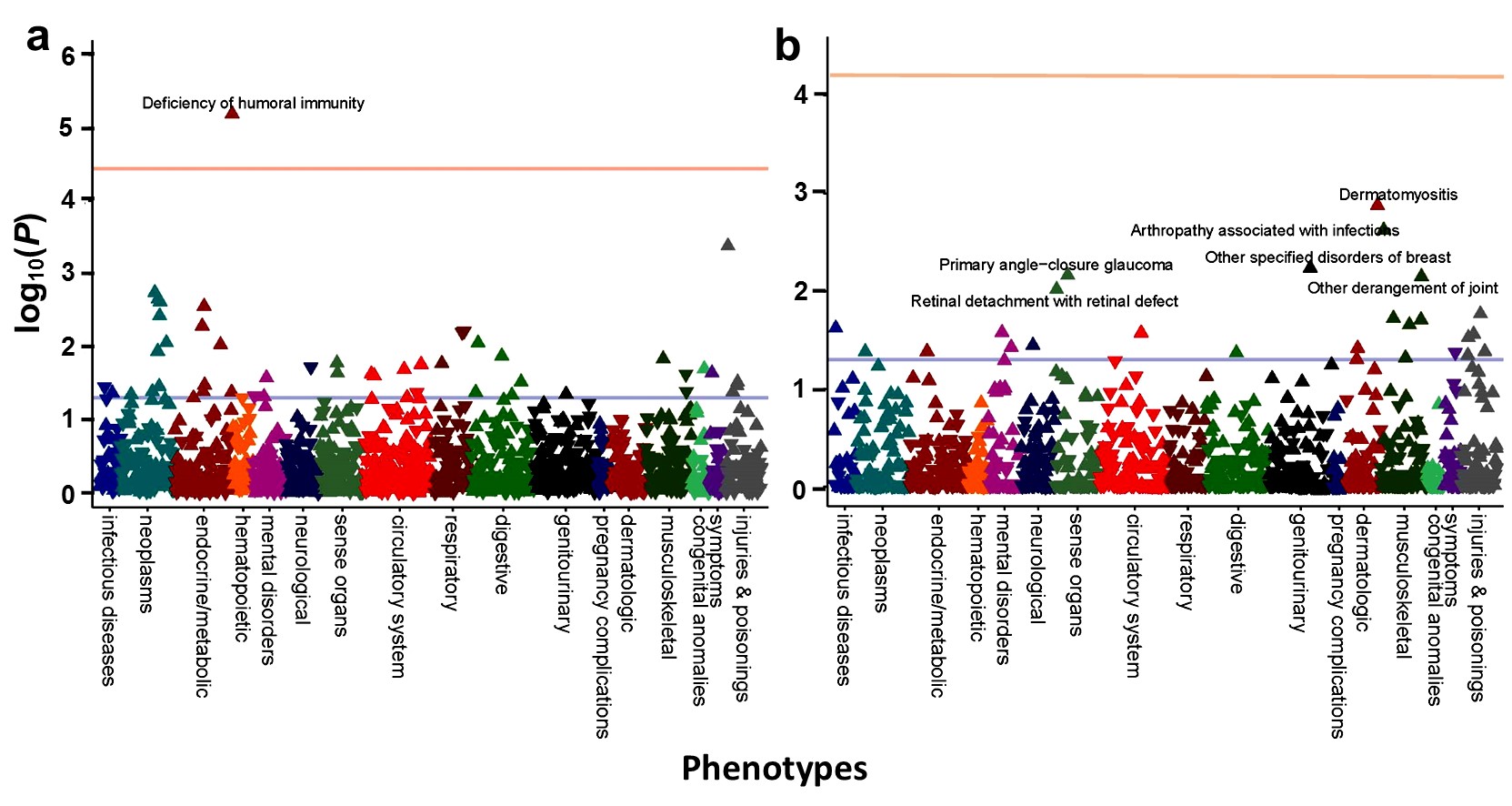

**Supplementary Figure 1. Recessive phenome-wide association study of rs1801725 (A986S) and rs1801726 (Q1011E) in the European descent dataset.** The Manhattan plot shows the phenotypes associated with (a) rs1801725 and (b) rs1801276 in an additive model. Upward arrows represent increased risk and downward arrows indicate decreased risk. None of the phenotypes passed Bonferroni correction (p < 3.65 x 10^-5^) represented by the orange line. The blue line represents p < 0.05.

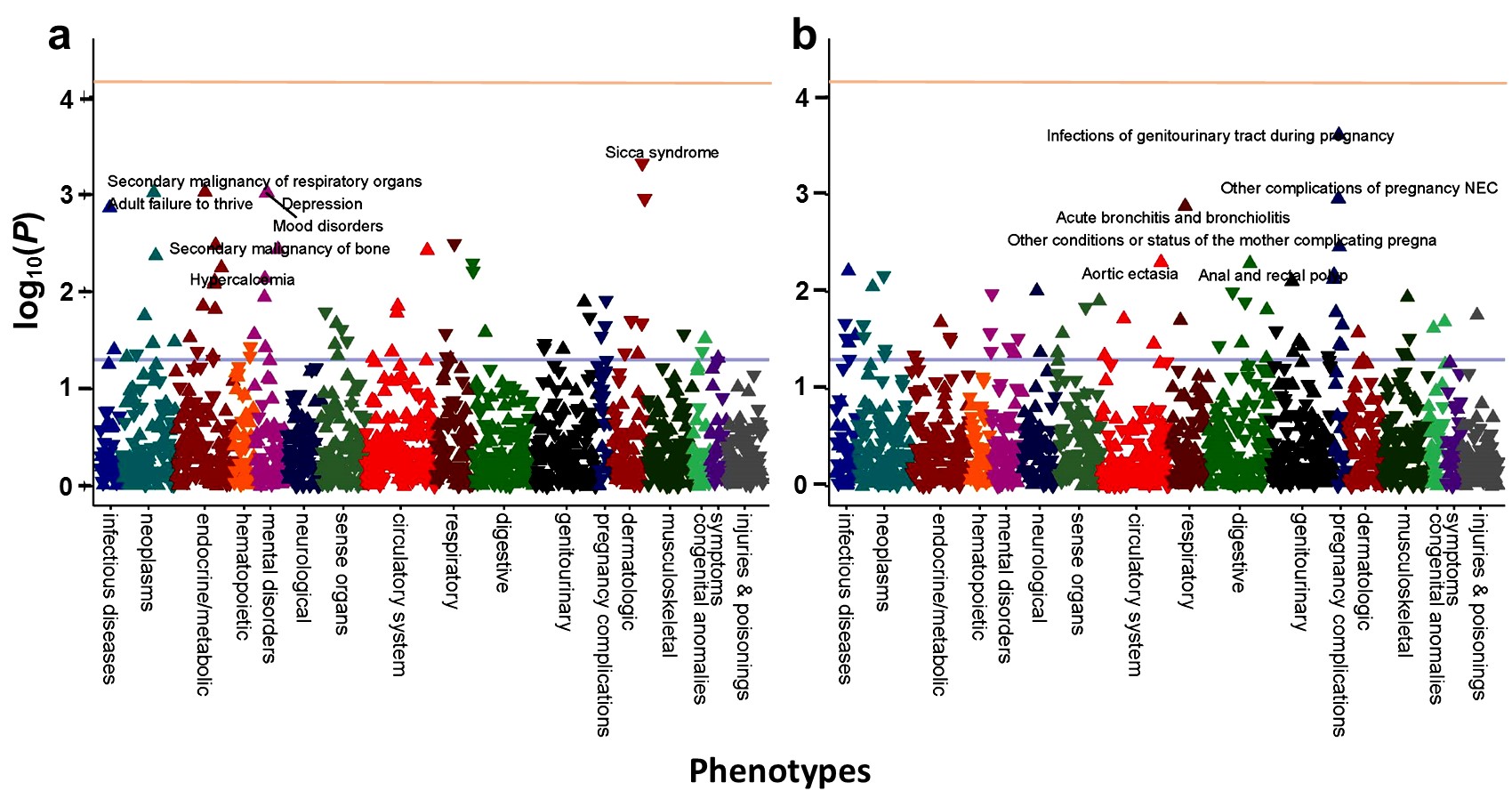

**Supplementary Figure 2. Additive phenome-wide association study of rs1801725 (A986S) and rs1801726 (Q1011E) in the European descent dataset.** The Manhattan plot shows the phenotypes associated with (a) rs1801725 and (b) rs1801276 in an additive model. Upward arrows represent increased risk and downward arrows indicate decreased risk. None of the phenotypes passed Bonferroni correction (p < 3.65 x 10^-5^) represented by the orange line. The blue line represents p < 0.05.

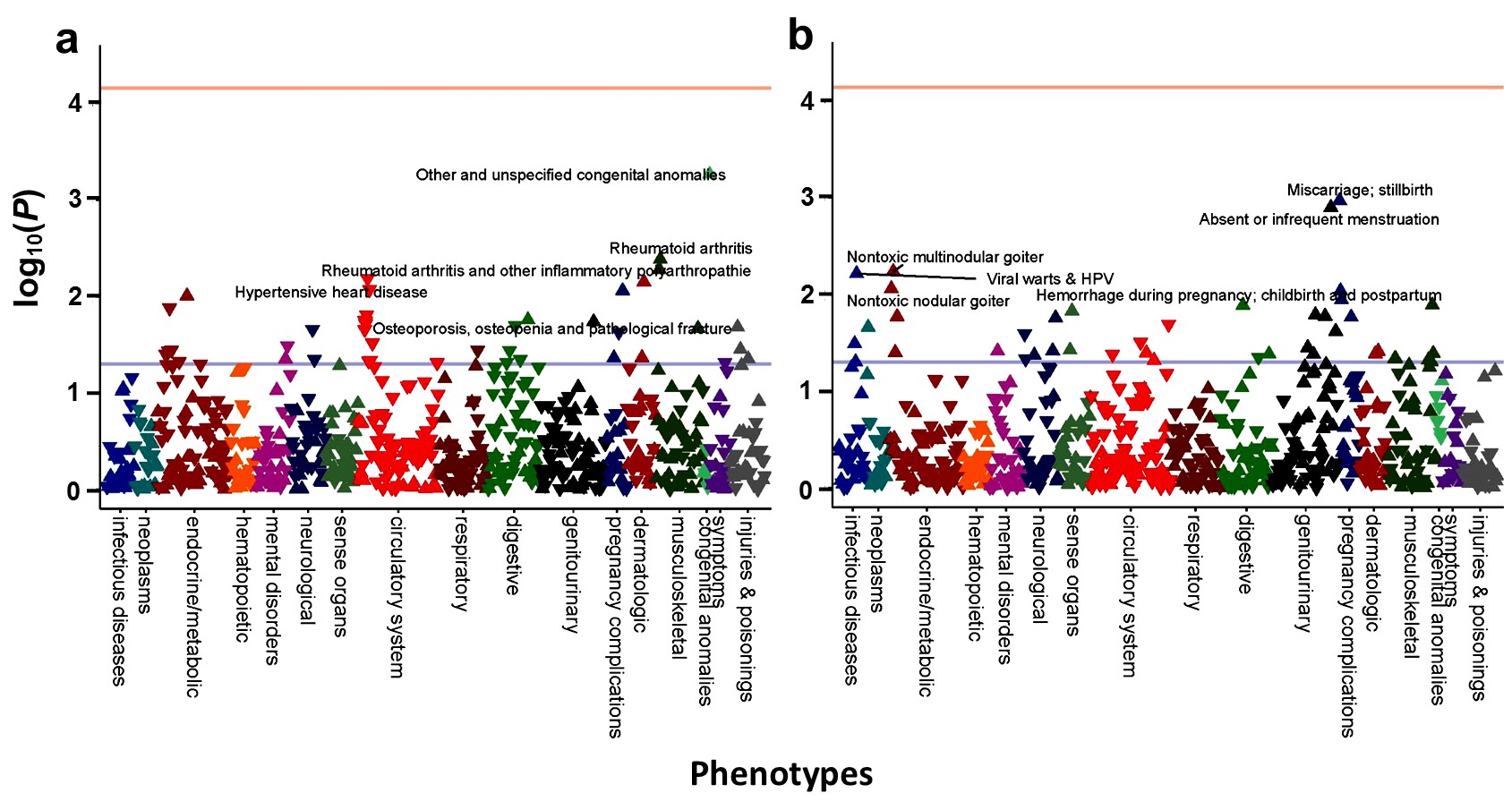

**Supplementary Figure 3. Recessive phenome-wide association study of rs1801725 (A986S) and rs1801726 (Q1011E) in the African descent dataset.** The Manhattan plot shows the phenotypes associated with (a) rs1801725 and (b) rs1801276 in an additive model. Upward arrows represent increased risk and downward arrows indicate decreased risk. None of the phenotypes passed Bonferroni correction (p < 7.33 x 10^-5^) represented by the orange line. The blue line represents p < 0.05.

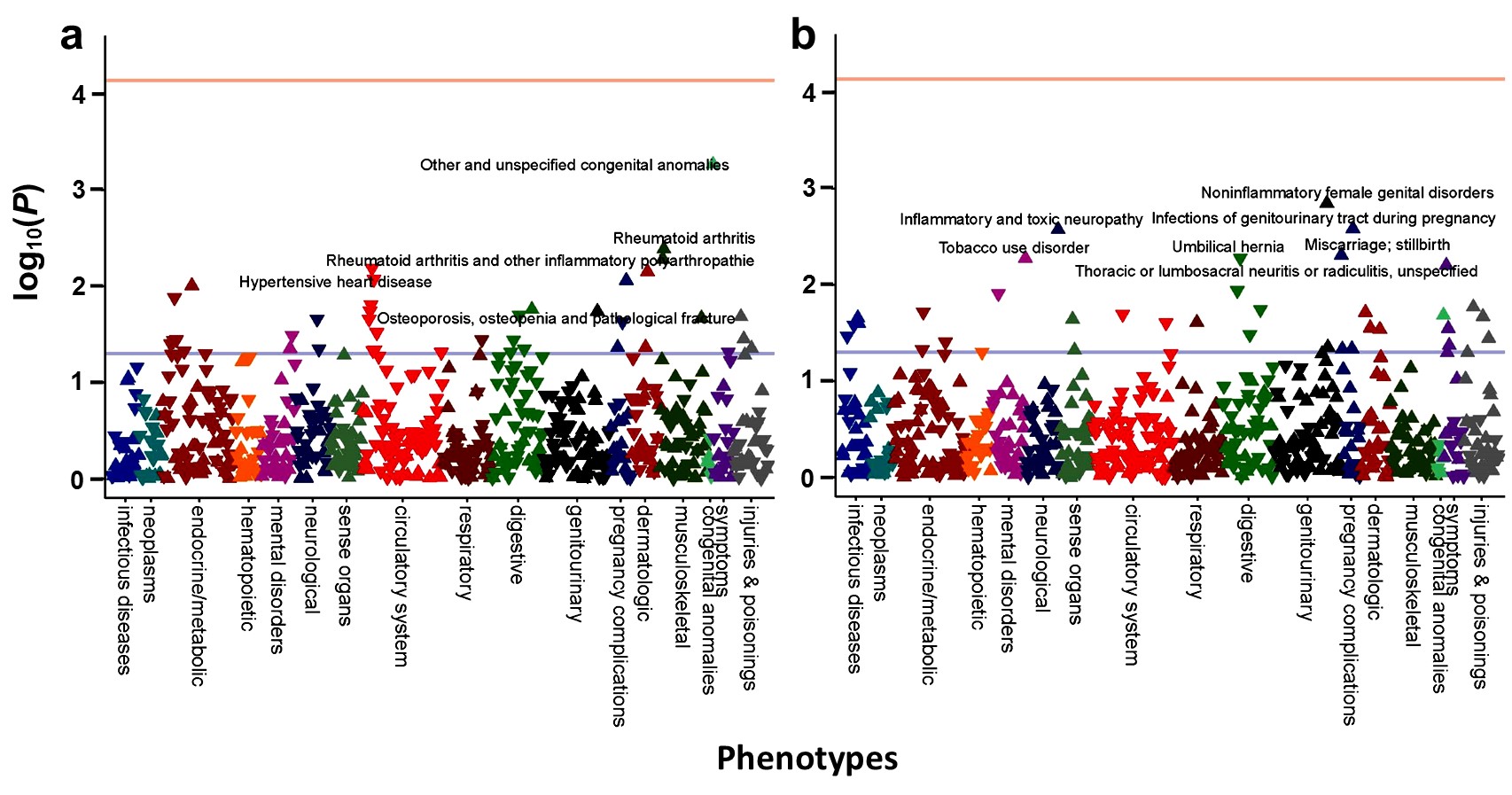

**Supplementary Figure 4. Additive phenome-wide association study of rs1801725 (A986S) and rs1801726 (Q1011E) in the African descent dataset.** The Manhattan plot shows the phenotypes associated with (a) rs1801725 and (b) rs1801276 in an additive model. Upward arrows represent increased risk and downward arrows indicate decreased risk. None of the phenotypes passed Bonferroni correction (p < 7.33 x 10^-5^) represented by the orange line. The blue line represents p < 0.05.
